# Face Masks Considerably Reduce Covid-19 Cases in Germany

**DOI:** 10.1101/2020.06.21.20128181

**Authors:** Timo Mitze, Reinhold Kosfeld, Johannes Rode, Klaus Wälde

## Abstract

We use the synthetic control method to analyze the effect of face masks on the spread of Covid-19 in Germany. Our identification approach exploits regional variation in the point in time when face masks became compulsory. Depending on the region we analyse, we find that face masks reduced the cumulative number of registered Covid-19 cases between 2.3% and 13% over a period of 10 days after they became compulsory. Assessing the credibility of the various estimates, we conclude that face masks reduce the daily growth rate of reported infections by around 40%.

## 1 Introduction

Many countries have experimented with several public health measures to mitigate the spread of Covid-19. One particular measure that has been introduced are face masks. It is of obvious interest to understand the contribution made by such a measure to reducing infections.

The effect of face masks on the spread of infections has been studied for a long time. The usefulness in the clinical context is beyond dispute. There is also considerable evidence that they helped in mitigating the spread of epidemics such as SARS 2003 or influenza (see below). The effect of face masks worn in public on the spread of Covid-19 has not been systematically analyzed so far. This is the objective of this paper.

There is a general perception in Germany that public wearing of face masks reduces incidences considerably. This perception comes mainly from the city of Jena. After face masks were introduced on 6 April 2020, the number of new infections fell almost to zero. Jena is not the only city or region in Germany, however, that introduced face masks. Face masks became compulsory in all federal states between 20 April and 29 April 2020. Six regions made masks compulsory before the introduction at the federal level. These dates lay between 6 April and 25 April (see appendix A and Kleyer et al., 2020, for a detailed overview of regulations in Germany). This leads to a lag between individual regions and the corresponding federal states of between two and 18 days.

We derive findings by employing synthetic control methods (SCM, Abadie and Gardeazabal, 2003, Abadie et al., 2010, Abadie, 2019). Our identification approach exploits the previously mentioned regional variation in the point in time when face masks became compulsory in public transport and sales shops. We use data for 401 German regions to estimate the effect of this public health measure on the development of registered infections with Covid-19. We consider the timing of mandatory face masks as an exogenous event to the local population. Masks were imposed by local authorities and were not the outcome of some process in which the population was involved.^2^ We compare the Covid-19 development in various regions to their synthetic counterparts. The latter are constructed as a weighted average of control regions that are similar to the regions of interest. Structural dimensions taken into account include prior Covid-19 cases, their demographic structure and the local health care system.

We indeed find strong and convincing statistical support for the general perception that public wearing of face masks in Jena strongly reduced the number of incidences. We obtain a synthetic control group that closely follows the Covid-19 trend before introduction of mandatory masks in Jena and the difference between Jena and this group is very large after 6 April. Our findings indicate that the early introduction of face masks in Jena has resulted in a reduction of almost 25% in the cumulative number of reported Covid-19 cases after 20 days. The drop is greatest, larger than 50%, for the age group 60 years and above. Our results are robust when we conduct sensitivity checks and apply several placebo tests, e.g. tests for pseudo-treatment effects in similarly sized cities in the federal state of Thuringia and for pseudo-treatment effects in Jena before the treatment actually started. We also test for announcement effects.

Constructing control groups for other single regions is not always as straightforward as for Jena. As a consequence, it is harder to identify the effect of face masks in these regions. When we move from single to multiple treatment effects, we find smaller effects. They are still sufficiently large, however, to support our point that wearing face masks is a very cost-efficient measure for fighting Covid-19. When we summarize all of our findings in one single measure (we compare all measures in appendix B.4), we conclude that the daily growth rate of Covid-19 cases in the synthetic control group falls by around 40% due to mandatory mask-wearing relative to the control group.^3^

Concerning the literature (see appendix D for a more detailed overview), the effects of face masks have been surveyed by Howard et al. (2020) and Greenhalgh et al. (2020). Greenhalgh et al. (2020) mainly presents evidence on the effect of face masks during non-Covid epidemics (influenza and SARS). Marasinghe (2020) reports that they “*did not find any studies that investigated the effectiveness of face mask use in limiting the spread of COVID-19 among those who are not medically diagnosed with COVID-19 to support current public health recommendations*”.

In addition to medical aspects (like transmission characteristics of Covid-19 and filtering capabilities of masks), Howard et al. (2020) survey evidence on mask efficiency and on the effect of a population. They first stress that “*no randomized control trials on the use of masks <…> has been published*”. The study which is “*the most relevant paper*” for Howard et al. (2020) is one that analyzed *“exhaled breath and coughs of children and adults with acute respiratory illness*” (Leung et al., 2020, p. 676), i.e. used a clinical setting. Concerning the effect of masks on community transmissions, the survey needs to rely on pre-Covid-19 studies. We conclude from this literature review that our paper is the first analysis that provides field evidence on the effect of masks on mitigating the spread of Covid-19.

## 2 Identification, data and implementation

### Identification

Our identification approach exploits the regional variation in the point in time when face masks became mandatory in public transport and sales shops. Given the federal structure of Germany, decisions are made by municipal districts (regions in what follows) and federal states. We can exploit differences by, first, identifying six regions (equivalent to the EU nomenclature of territorial units for statistics, NUTS, level 3) which made wearing face masks compulsory before their respective federal states. For all other regions, mandatory mask-wearing followed the decision of the corresponding federal state. Second, as Figure 1 shows, variation across federal states also implies variations across regions.

**Figure 1:**
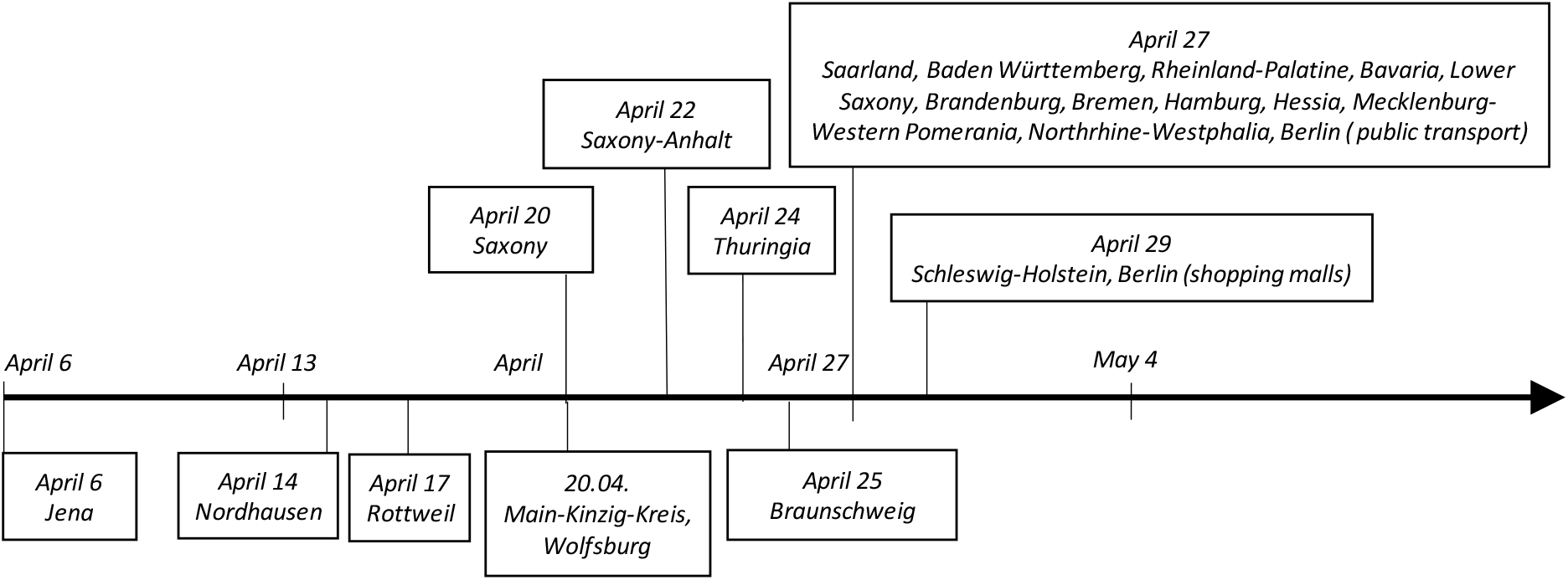
The timing of mandatory mask wearing in federal states (top) and individual regions (below)

To identify possible treatment effects from introducing face masks, we apply SCM for single and multiple treated units. Our methodical choice is motivated as follows: First, the original goal of SCM to “*estimate the effects of <…> interventions that are implemented at an aggregate level affecting a small number of large units (such as cities, regions, or countries)*” (Abadie, 2019, p.3) clearly matches with our empirical setup. Compared to standard regression analyses, SCM is particularly well suited for comparative case study analyses with only one treated unit or a very small number thereof (Abadie and Gardeazabal, 2003, Becker et al., 2018). Second, the method is flexible, transparent and has become a widely utilized tool in the policy evaluation literature (Athey and Imbens, 2017) and for causal analyses in related disciplines (see, e.g., Kreif et al., 2015, for an overview of SCM in health economics, Pieters et al., 2017, for a biomedical application).^4^

SCM identifies synthetic control groups for the treated unit(s) to build a counterfactual. In our case, we need to find a group of regions that have followed the same Covid-19 trend as treated units before mandatory masks in the latter. This control group would then most likely have had the same behavior as treated unit(s) in the absence of the mask obligation. We can then use this group to ‘synthesize’ the treated unit and conduct causal inference. The synthetic control group is thereby constructed as an estimated weighted average of all regions in which masks did not become compulsory earlier on. Historical realizations of the outcome variable and several other predictor variables that are relevant in determining outcome levels allow us to generate the associated weights, which result from minimizing a pre-treatment prediction error function (see Abadie and Gardeazabal, 2003, Abadie et al., 2010 and Abadie, 2019 for methodical details).

### Data

We use the official German statistics on reported Covid-19 cases from the Robert Koch Institute (RKI, 2020). The RKI collects the data from local health authorities and provides updates on a daily basis. Using these data (available via API), we build a balanced panel for 401 NUTS Level 3 regions and 95 days spanning the period from January 28 to May 1, 2020 (38,095 observations). We use the cumulative number of registered Covid-19 cases in each district as main outcome variable.^5^ We estimate overall effects for this variable together with disaggregated effects by age groups (persons aged 15-34 years, 35-59 years and 60+ years). As an alternative outcome variable, we also use the cumulative incidence rate. Table 1 shows summary statistics for both variables for our sample period.

**Table 1:** Summary Statistics of Covid-19 indicators (outcome variables) and predictors characterizing the regional demographic structure and basic health care system

|  | Mean | S.D. | Min. | Max. |
| --- | --- | --- | --- | --- |
| **PANEL A: Data on registered Covid-19 cases** |  |  |  |  |
| [1] Newly registered cases per day | 4.13 | 10.66 | 0 | 310 |
| [2] Cumulative number of cases | 120.86 | 289.07 | 0 | 5795 |
| [3] Cum. cases [2] per 100,000 inhabitants | 59.87 | 106.80 | 0 | 1,530.32 |
| **PANEL B: Regional demographic structure and local health care system** |  |  |  |  |
| Population density (inhabitants/km<sup>2</sup>) | 534.79 | 702.40 | 36.13 | 4,686.17 |
| Population share of highly educated* individuals (in %) | 13.07 | 6.20 | 5.59 | 42.93 |
| Share of females in population (in %) | 50.59 | 0.64 | 48.39 | 52.74 |
| Average age of females in population (in years) | 45.86 | 2.11 | 40.70 | 52.12 |
| Average age of males in population (in years) | 43.17 | 1.83 | 38.80 | 48.20 |
| Old-age dependency ratio (persons aged 65 years and above per 100 of population age 15-64) | 34.34 | 5.46 | 22.40 | 53.98 |
| Young-age dependency ratio (persons aged 14 years and below per 100 of population age 15-64) | 20.54 | 1.44 | 15.08 | 24.68 |
| Physicians per 10,000 of population | 14.58 | 4.41 | 7.33 | 30.48 |
| Pharmacies per 100,000 of population | 27.01 | 4.90 | 18.15 | 51.68 |
| Settlement type (categorical variable<sup>§</sup>) | 2.59 | 1.04 | 1 | 4 |

Notes: \* = International Standard Classification of Education (ISCED) Level 6 and above; § = categories are based on population shares and comprise 1) district-free cities (*kreisfreie Großstädte*), 2) urban districts (*städtische Kreise*), 3) rural districts (*ländliche Kreise mit Verdichtungsansätzen*), 4) sparsely populated rural districts (*dünn besiedelte ländliche Kreise*).

Table 1 also presents our other predictor variables. We focus on factors that are likely to describe the regional number and dynamics of reported Covid-19 cases. Obviously, past values of (newly) registered Covid-19 cases are important to predict the regional evolution of Covid-19 cases over time in an autoregressive manner. In addition, we argue that a region’s demographic structure, such as the overall population density and age structure, and its basic health care system, such as the regional endowment with physicians and pharmacies per population, are important factors for characterizing the local context of Covid-19. Predictor variables are obtained from the INKAR online database of the Federal Institute for Research on Building, Urban Affairs and Spatial Development. We use the latest year available in the database (2017). We consider it likely that regional demographic structures only gradually vary over time such that they can be used to measure regional differences during the spread of Covid-19 in early 2020.

### Implementation

The implementation of the SCM is organized as follows: As baseline analysis, we focus on the single treatment case for the city of Jena for three reasons. First, as shown in Figure 1, Jena was the first region to introduce face masks in public transport and sales shops on April 6. This results in a lead time of 18 days relative to mandatory face masks in the surrounding federal state Thuringia on April 24. By April 29, all German regions had introduced face masks (exact dates are provided in appendix A). A sufficiently long lag between mandatory face masks in the treated unit vis-à-vis the sample of control regions is important for effect identification.

Second, the timing of the introduction of face masks in Jena is -by and large-not affected by other overlapping public health measures related to the Covid-19 spread. Since March 22 the German economy had been in a general “lock down” coordinated among all federal states. Only from April 20 onwards has the economy been gradually reopening. Third, Jena is in various ways a representative case for studying the Covid-19 development: On April 5, which is one day before face masks became compulsory in Jena, the cumulative number of registered Covid-19 cases in Jena was 144. This is very close to the median of 155 for Germany. Similarly, the cumulative number of Covid-19 incidences per 100,000 inhabitants was 126.9 in Jena compared to a mean of 119.3 in Germany (compare Figure A1).

In our baseline configuration of the SCM, we construct the synthetic Jena by including the number of cumulative Covid-19 cases (measured one and seven days before the start of the treatment) and the number of newly registered Covid-19 cases (in the last seven days prior to the start of the treatment) as autoregressive predictor variables. The chosen period shall ensure that the highly non-linear short-run dynamics of regional Covid-19 cases are properly captured. We use cross-validation tests to check the sensitivity of the SCM results when we allow for a shorter training period in the pre-treatment phase by imposing longer lags. The autoregressive predictors are complemented by the cross-sectional data on the region’s demographic and basic health care structure.

Although the case study of Jena can be framed in a clear identification strategy, the Covid-19 spread in a single municipality may still be driven by certain particularities and random events that may prevent a generalization of estimated effects. We therefore also test for treatment effect in districts that introduced face masks after Jena but still before they became compulsory in the corresponding federal state. More importantly, however, we apply a multiple treatment approach that takes all regions as treated units which introduced face masks by April 22. This results in 32 regions from Saxony and Saxony-Anhalt. All other regions apart from Thuringia introduced face masks on April 27. We employ this delay to study the effects of mandatory masks up to May 1^st^. We end on May 1^st^ as we would expect that differences across treated and non-treated regions should disappear 5-7 days after April 27. This delay results from a median incubation time of 5.2 days (Linton et al., 2020 and Lauer et al., 2020) and around 2 days accounting for reporting to authorities (as assumed e.g. in Donsimoni et al., 2020a, b).

Although SCM appears to be a natural choice for our empirical identification strategy, we are well aware of the fact that its validity crucially depends on important practical requirements including the availability of a proper comparison group, the absence of early anticipation effects or interference from other events (Cavallo et al., 2013, Abadie, 2019). In the implementation of the single and multiple treatment SCM we check for these pitfalls through sensitivity and placebo tests. We deal with these issues in our baseline case study for Jena as follows:

1. We have screened the introduction and easing of public health measures, as documented in Kleyer et al. (2020), to ensure that no interference takes place during our period of study. This is the case at least until April 20 when exit strategies from public health measures started.
2. We make sure that the regions used to create the synthetic control, i.e. the donor pool, are not affected by the treatment (Campos et al., 2015). We eliminate the two immediate geographical neighbors of Jena from the donor pool to rule out spillover effects. We also exclude those regions for which anticipation effects may have taken place because face masks became compulsory in quick succession to Jena.
3. We account for early anticipation effects in Jena. Specifically, we take the announcement that face masks will become compulsory one week before their introduction as an alternative start of the treatment period.
4. We apply cross-validation tests to check for sensitivities related to changes in historical values in the outcome variables used as predictors. We also run placebo-in-time tests to check whether effects actually occurred even before the start of the treatment.
5. We test for the sensitivity of the results when changing the donor pool and run comprehensive placebo-in-space tests as a mode of inference in the SCM framework.

Inference thereby relies on permutation tests and follows the procedures suggested by Cavallo et al. (2013) and applied, for example, by Eliason and Lutz (2018) or Hu et al. (2018). For both the single and multiple treatment applications we estimate placebo-treatment effects for each district in which masks did not become compulsory early on. These placebo treatments should be small, relative to the treated regions. We calculate significance levels for the test of the hypothesis that the mask obligation did not significantly affect reported Covid-19 cases. This provides us with *p*−values for each day, which capture the estimated treatment effect on reported Covid-19 cases from placebo regions. The *p*-values are derived from a ranking of the actual treatment effect within the distribution of placebo treatment effects. We follow the suggestion in Galiani and Quistorff (2017) and compute adjusted *p*-values taking the pre-treatment match quality of the placebo treatments into account.^6^

## 3 The effects of face masks on Covid-19

### Baseline results for Jena

Panel A in Figure 2 shows the SCM results for the introduction of face masks in Jena on April 6. The visual inspection of the development of cumulative Covid-19 cases shows that the fit of the synthetic control group is very similar to Jena before the treatment.^7^ The difference in the cumulated registered Covid-19 cases between Jena and its corresponding synthetic control unit after the start of the treatment can be interpreted as the treatment effect on the treated.

**Figure 2:**
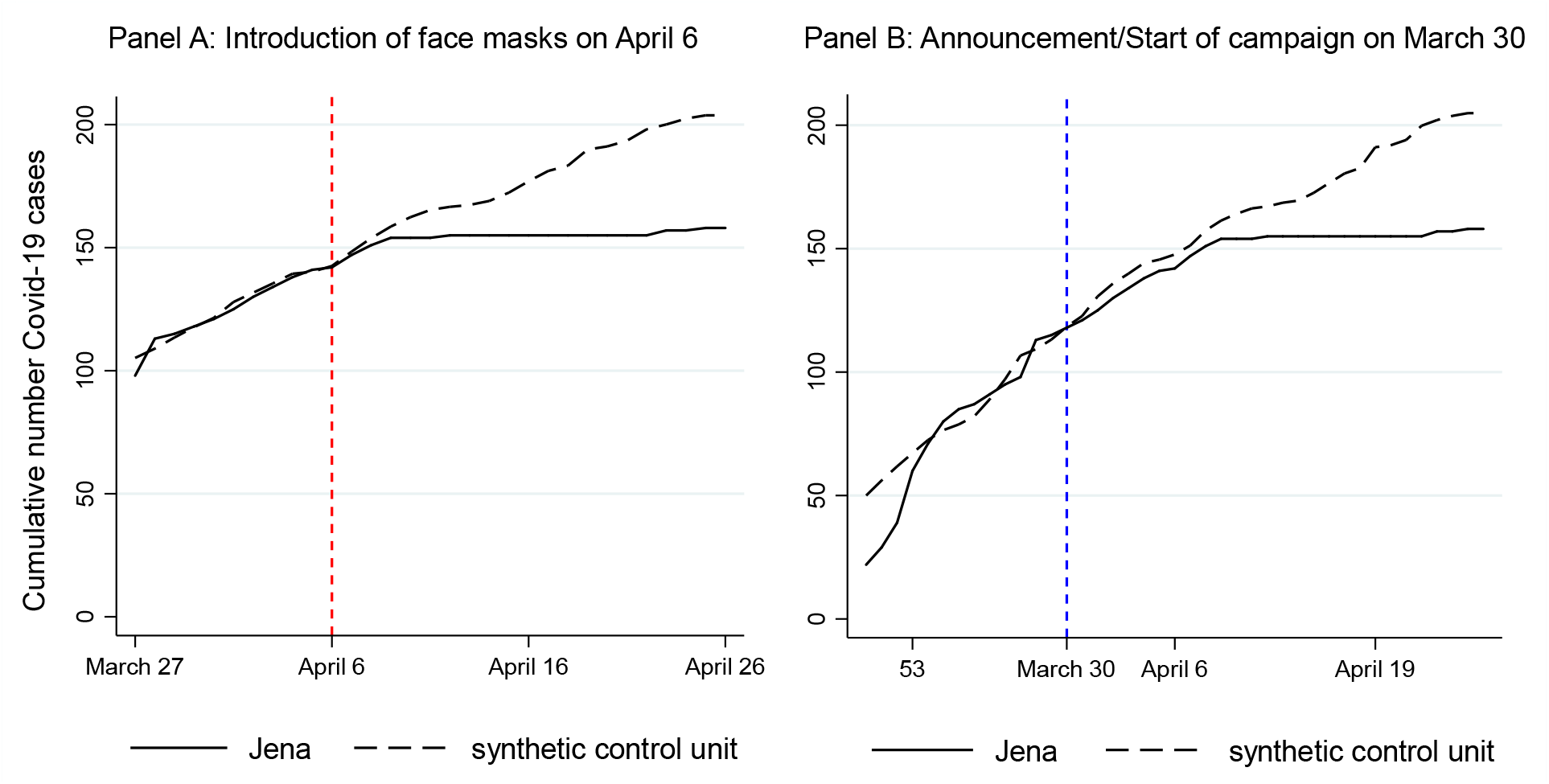
Treatment effects of mandatory face masks in Jena on April 6 and start of campaign on March 30 (see Table A3 and appendix B.2 for details)

The figure clearly shows a gradually widening gap in the cumulative number of Covid-19 cases between Jena and the synthetic control unit. The size of the effect 20 days after the start of the treatment (April 26) amounts to a decrease in the number of cumulative Covid-19 cases of 23%. For the first 10 days, the decrease amounts to 13%. Expressed differently, the daily growth rate of the number of infections decreases by 1.32 percentage points per day (see appendix B.4 for computational details and an overview of all measures). If we look at the estimated differences by age groups, Table A2 in the appendix indicates that the largest effects are due to the age group of persons aged 60 years and above. Here the reduction in the number of registered cases is even larger than 50%. For the other two age groups we find a decrease between 10 and 20%.

If we consider a median incubation of 5.2 days plus a potential testing and reporting lag of 2-3 days, the occurrence of a gradually widening gap between Jena and its synthetic control three to four days after the mandatory face masks seems fast. One might conjecture that an announcement effect played a role. As shown in appendix B.7, online searches for (purchasing) face masks peaked on April 22, when it was announced that face masks would become compulsory in all German federal states.^8^ A smaller peak (70% of the April 22 peak) of online searches appeared on March 31. This is one day after Jena announced that masks would become compulsory on April 6. The announcement was accompanied by a campaign “Jena zeigt Maske” to communicate the necessity to wear face masks in public^9^ and was widely discussed all over Germany.

Panel B in Figure 2 therefore plots the results when we set the start of the treatment period to the day of the announcement on 30 March. The visual inspection of the figure shows the existence of a small anticipation effect (which is mainly driven by the relative development of Covid-19 age group 15-34 years (Panel B in Figure A2). Yet, the gap to the synthetic control significantly widens only approximately 10 days after the announcement. As this temporal transmission channel appears plausible against the background of incubation times and given that no other intervention took place around this time in Jena or the regions in the synthetic control group, we take this as first evidence for a face mask-effect in the reduction of Covid-19 infections. Appendix B.6 shows similar SCM results for the incidence rate (overall and by age groups). We find a reduction of approximately 30 cases per 100,000 of population.

Obviously, the estimated differences in the development of Jena vis-à-vis the synthetic Jena is only consistently estimated if our SCM approach delivers robust results. Accordingly, we have applied several tests to check for the sensitivity of our findings.

### Cross-validation and placebo-in-time test

One important factor is that our results are not sensitive to changes in predictor variables. We therefore perform cross-validation checks by modifying the length of the training and validation periods before the start of the treatment. Panel A in Figure 3 shows that lagging the autoregressive predictor variables further in time only slightly changes our results. Importantly, we do not find a systematic downward bias of our baseline specification (cumulative number of reported Covid-19 cases: one and seven days before start of treatment; number of newly registered Covid-19 cases: last seven days before start of treatment) compared to an alternative specification. The latter trains the synthetic control on the basis of information on cumulative Covid-19 cases 7 and 14 days prior to the treatment together with the development of newly register cases between day 7 and 14 prior to the treatment. Given that regional Covid-19 cases developed very dynamically and non-linearly in this period, this is an important finding in terms of the robustness of our results.

**Figure 3:**
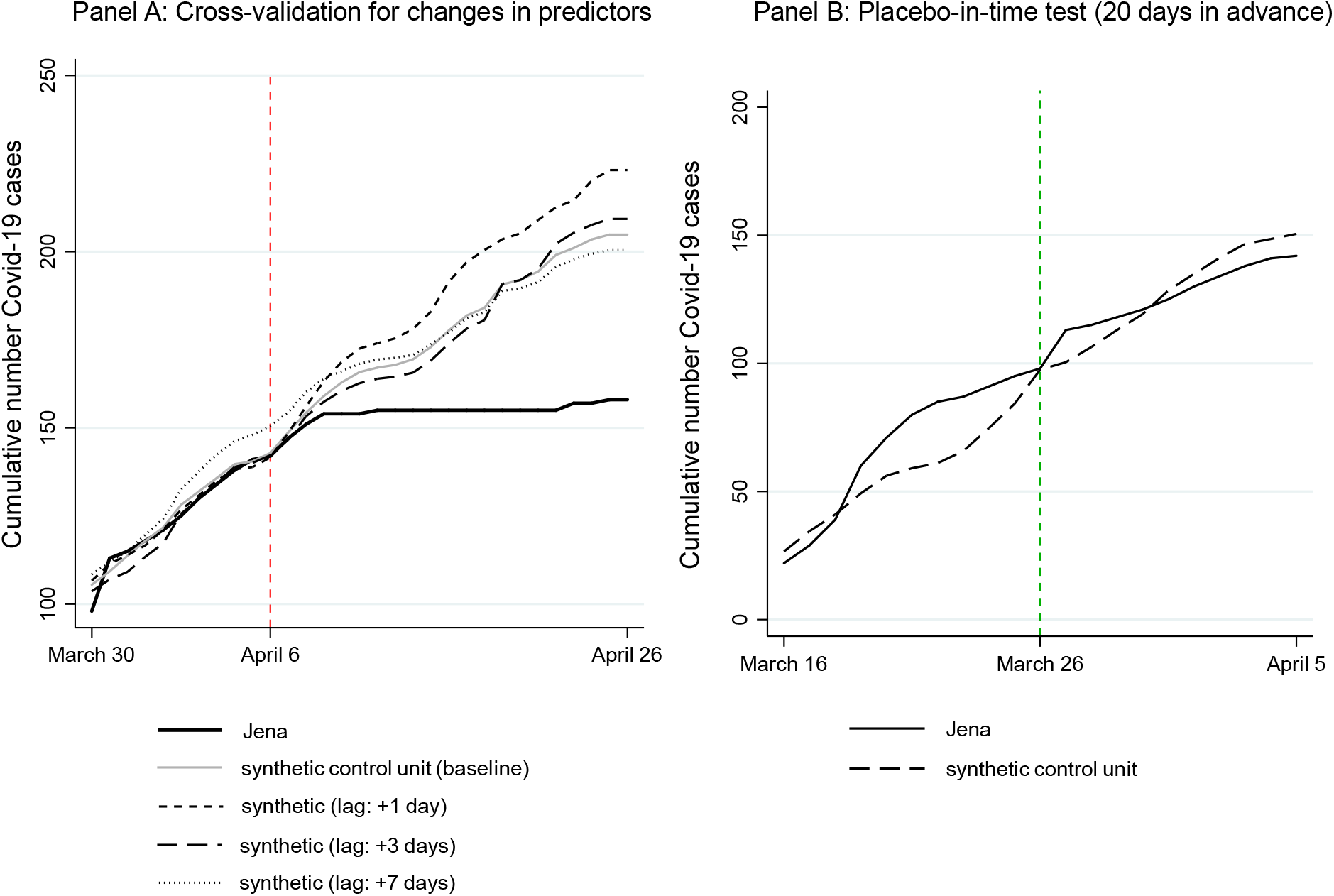
Cross-validation for changes in predictor variables and placebo-in-time test *Notes:* In Panel A the baseline specification for the synthetic control group uses historical values of the outcome variable in the following way: i) number of cumulative Covid-19 cases (measured one and seven days before the start of the treatment), ii) the number of newly registered Covid-19 cases (in the last seven days prior to the start of the treatment); the alternative specifications lag these values by 1, 3 and 7 days.

Another important factor for the validity of the results is that we do not observe an anticipation effect for Jena prior to the announcement day. We test for a pseudo-treatment in Jena over a period of 20 days before the introduction of face masks. This period is equally split into a pre- and pseudo post-treatment period. As Panel B in Figure 3 shows, there is no strong deviation from the path of the synthetic control group. This result needs to be interpreted with some care as the regional variation of Covid-19 cases in Germany is very heterogeneous the longer we go back in time. This is indicated by the generally lower fit of the synthetic control group in matching the development in Jena in mid-March when the absolute number of Covid-19 cases was still low.

### Changing the donor pool

In addition, we also check for the sensitivity of the results when changing the donor pool. This may be important as our baseline specification includes the region of Heinsberg in the donor pool used to construct the synthetic Jena (with a weight of 4.6%; compare Table A3). As Heinsberg is one of the German regions which was significantly affected by the Covid-19 pandemic during the Carnival season, this may lead to an overestimation of the effects of face masks. Accordingly, appendix B.8 presents estimates for alternative donor pools. Again, we do not find evidence for a significant bias in our baseline specification. By tendency, the treatment effect becomes larger, particularly if we compare Jena only to other regions in Thuringia (to rule out macro-regional trends) and to a subsample of larger cities (*kreisfreie Städte*). Both subsamples exclude Heinsberg. We also run SCM for subsamples excluding Thuringia (to rule out spillover effects) and for East and West Germany (again in search for specific macro regional trends). Generally, these sensitivity tests underline the robustness of the estimated treatment effect for Jena.

### Placebo-in-space tests

A placebo test in space checks whether other cities that did not introduce face masks on April 6 have nonetheless experienced a decline in the number of registered Covid-19 cases. If this had been the case, the treatment effect may be driven by other latent factors rather than face masks. Such latent factors may, for instance, be related to the macro-regional dynamics of Covid-19 in Germany. Therefore, appendix B.9 reports pseudo-treatment effects for similarly sized cities in Thuringia assuming that they have introduced face masks on April 6 although −in fact− they did not. As the figure shows, these cities show either a significantly higher or similar development of registered Covid-19 compared to their synthetic controls. This result provides further empirical support for a relevant effect in the case of Jena.

As a more comprehensive test, we also ran placebo-in-space tests for all other regions that did not introduce face masks on April 6 or closely afterwards. Again, we estimate the same model on each untreated region, assuming it was treated at the same time as Jena. The empirical results in Figure 4 indicate that the reduction in the reported number of Covid-19 cases in Jena clearly exceeds the trend in most other regions − both for the overall sample in Panel A and the subsample of large cities (*kreisfreie Städte*) in Panel B.

**Figure 4:**
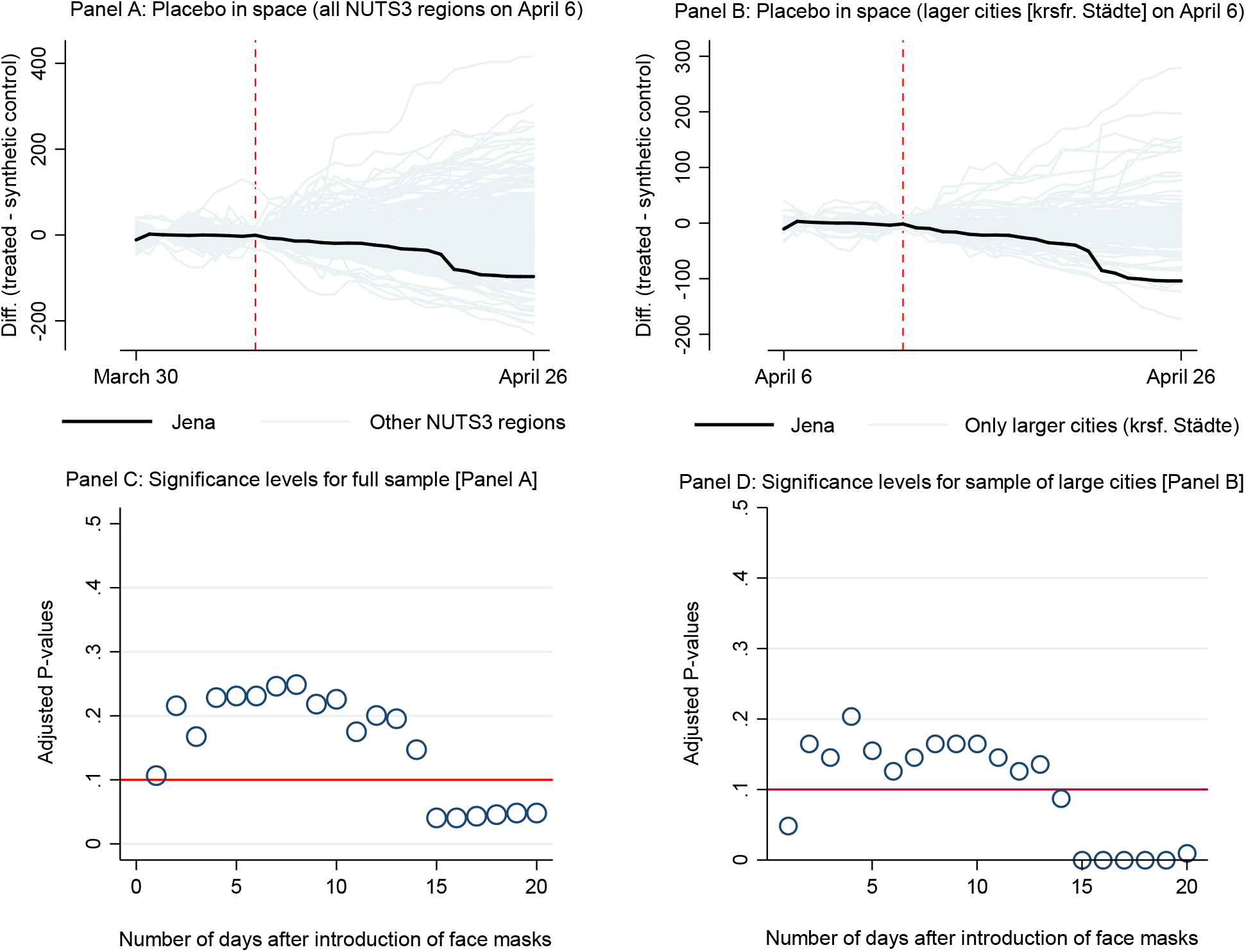
Comprehensive placebo-in-space tests for the effect of face masks on Covid-19 cases *Notes:* Graphs exclude the following regions with a very large number of registered Covid-19 cases: Hamburg (2000), Berlin (11000), Munich (9162), Cologne (5315) and Heinsberg (5370). In line with Abadie et al. (2010), we only include placebo effects in the pool for inference if the match quality (pre-treatment RMSPE) of the specific control regions is smaller than 20 times the match quality of the treated unit. *P*-values are adjusted for the quality of the pre-treatment matches (see Galiani and Quistorff, 2017).

As outlined above, one advantage of this type of placebo-in-space-test is allows us to conduct inference. Accordingly, Panel C and Panel D report adjusted *p*-values that indicate the probability if the treatment effect for Jena was observed by chance given the distribution of pseudo-treatment effects in the other German regions (see Galiani and Quistorff, 2017). For both samples, the reported *p*-values indicate that the reduction in the number of Covid-19 cases in Jena did not happen by chance but can be attributed to the introduction of face masks, at the latest - roughly two weeks after the start of the treatment. This timing is again in line with our above argument that a sufficiently long incubation time and testing lags need to be considered in the evaluation of treatment effects.

### Treatment in other districts

Jena may be a unique case. We therefore also study treatment effects for other regions that have antedated the general introduction of face masks in Germany. Further single unit treatment analyses are shown in appendix C. Multiple unit treatments are studied in two ways. The first sample covers all 401 regions and 32 treated units. The second focused on the subsample of 105 larger cities (*kreisfreie Städte*), of which 8 are treated units. Treated regions introduced face masks by April 22. The multiple treatment approach, visible in Figure 5, points to a significant face mask-effect in the reduction of Covid-19 infections. The adjusted *p*-values indicate that the estimated treatment effects are not random.

**Figure 5:**
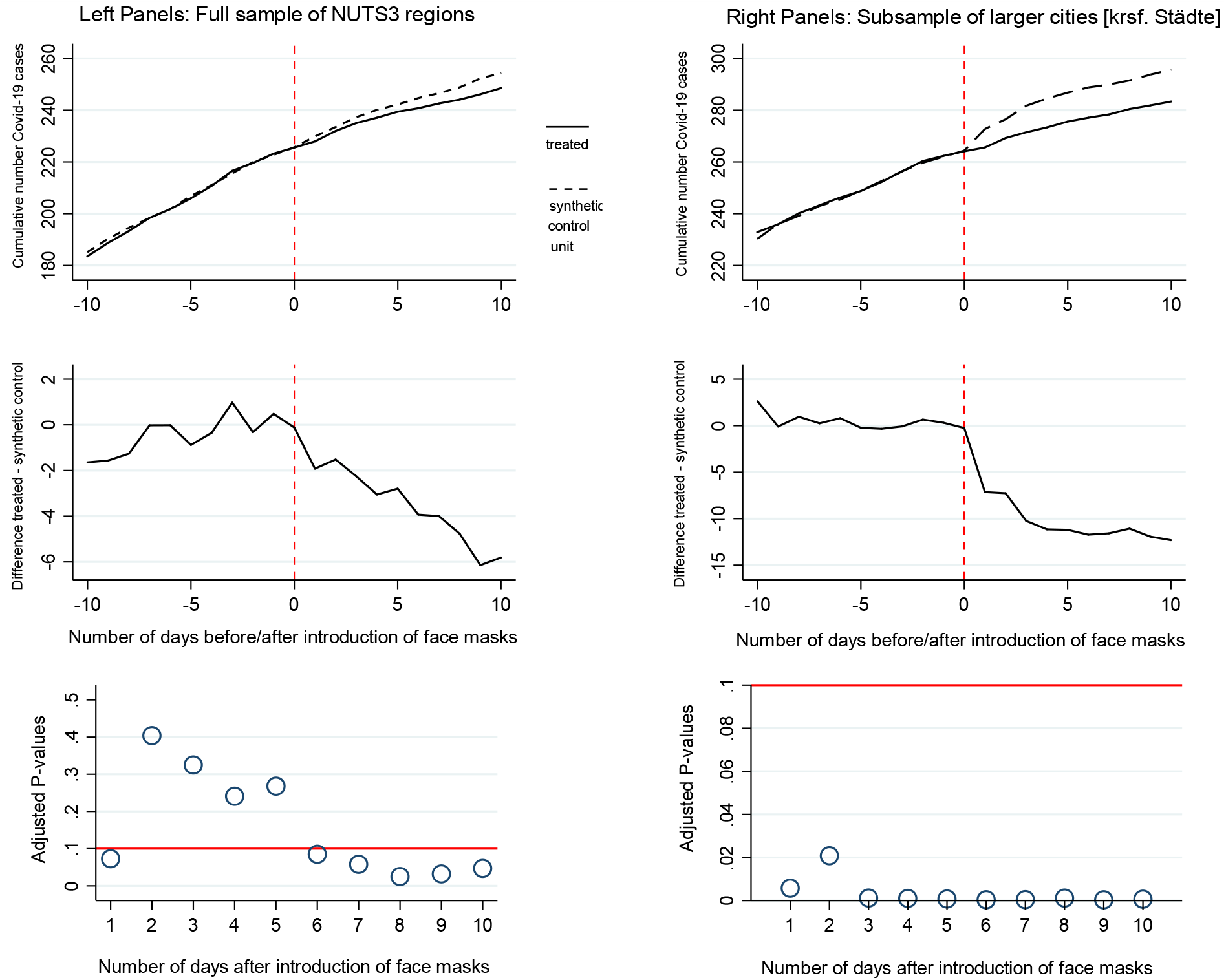
Average treatment effects for introduction of face masks (multiple treated units) *Notes* Statistical inference for adjusted *p*-values has been conducted on the basis of a random sample of 1,000,000 placebo averages.

Face masks may have made a particular difference in the spread of Covid-19, particularly in larger cities with higher population density and accordingly higher intensity of social interaction.^10^ Over a period of 10 days, we observe an average reduction of 12.3 cases between treated and control regions. Relative to the average number of cumulative Covid-19 cases on May 1 in control regions (295.6), this amounts to a reduction of 4.2% of cases. The daily growth rate of the number of infections correspondingly shrinks by 0.42 percentage points. For the entire sample, the reduction in the daily growth rate is estimated to be 0.23 percentage points (see again appendix B.4 for an overview of all measures).

## 4 Conclusion

We set out by analyzing the city of Jena. The introduction of face masks on 6 April reduced the number of new infections over the next 20 days by almost 25% relative to the synthetic control group. This corresponds to a reduction in the average daily growth rate of the total number of reported infections by 1.32 percentage points. Comparing the daily growth rate in the synthetic control group with the observed daily growth rate in Jena, the latter shrinks by around 60% due to the introduction of face masks. This is a sizeable effect. Wearing face masks apparently helped considerably in reducing the spread of Covid-19. Looking at single treatment effects for all other regions puts this result in some perspective. The reduction in the growth rate of infections amounts to 20% only. By contrast, when we take the multiple treatment effect for larger cities into account, we find a reduction in the growth rate of infections by around 40%. What would we reply if we were asked what the effect of introducing face masks would have been if they had been made compulsory all over Germany? The answer depends, first, on which of the three percentage measures we found above is the most convincing and, second, on the point in time when face masks are made compulsory. The second aspect is definitely not only of academic interest but would play a major role in the case of a second wave.

We believe that the reduction in the growth rates of infections by 40% to 60% is our best estimate of the effects of face masks. The most convincing argument stresses that Jena introduced face masks before any other region did so. It announced face masks as the first region in Germany while in our post-treatment period no other public health measures were introduced or eased. Hence, it provides the most clear-cut experiment of its effects. Second, as stated above, Jena is a fairly representative region of Germany in terms of Covid-19 cases. Third, the smaller effects observed in the multiple treatment analysis may also result from the fact that −by the time that other regions followed the example of Jena− behavioral adjustments in Germany’s population had also taken place. Wearing face masks gradually became more common and more and more people started to adopt their usage even when it was not yet required.

We should also stress that 40 to 60% might still be a lower bound. The daily growth rates in the number of infections when face masks were introduced was around 2 to 3%. These are very low growth rates compared to the early days of the epidemic in Germany, where daily growth rates also lay above 50% (Wälde, 2020). One might therefore conjecture that the effects might have been even greater if masks had been introduced earlier.

We simultaneously stress the need for more detailed analyses. First, Germany is only one country. Different norms or climatic conditions might change the picture for other countries. Second, we have ignored spatial dependencies in the epidemic diffusion of Covid-19. This might play a role. Third, there are various types of face masks. We cannot identify differential effects since mask regulations in German regions do not require a certain type. This calls for further systematic causal analyses of the different health measure implemented to fight the spread of Covid-19. Our results provide some initial empirical evidence on this important matter.

## Data Availability

We use the official German statistics on reported Covid-19 cases from the Robert Koch Institute (RKI, 2020). The RKI collects the data from local health authorities and provides updates on a daily basis. Using these data (available via API), we build a balanced panel for 401 NUTS Level 3 regions and 95 days spanning the period from January 28 to May 1, 2020 (38,095 observations). We use the cumulative number of registered Covid-19 cases in each district as main outcome variable. We estimate overall effects for this variable together with disaggregated effects by age groups (persons aged 15-34 years, 35-59 years and 60+ years). As an alternative outcome variable, we also use the cumulative incidence rate. Table 1 shows summary statistics for both variables for our sample period.

https://npgeo-corona-npgeo-de.hub.arcgis.com/

## Supplementary Appendix for

### A. Timing of introduction of mandatory face masks

**Table A1:** Overview of dates when masks became compulsory in federal states and districts

| **Federal State** | **Public transport** | **Sales shops** | **Individual NUTS3 region** | **Introduction of face masks** | **Difference in days to state** |
| --- | --- | --- | --- | --- | --- |
| Baden-Wuerttemberg | 27.04.2020 | 27.04.2020 | LK Rottweil | 17.04.2020 | 10 |
| Bavaria | 27.04.2020 | 27.04.2020 |  |  |  |
| Berlin | 27.04.2020 | 29.04.2020 |  |  |  |
| Brandenburg | 27.04.2020 | 27.04.2020 |  |  |  |
| Bremen | 27.04.2020 | 27.04.2020 |  |  |  |
| Hamburg | 27.04.2020 | 27.04.2020 |  |  |  |
| Hesse | 27.04.2020 | 27.04.2020 | Main-Kinzig-Kreis | 20.04.2020 | 7 |
| Mecklenburg-West Pomer. | 27.04.2020 | 27.04.2020 |  |  |  |
| Lower Saxony | 27.04.2020 | 27.04.2020 | Wolfsburg | 20.04.2020 | 7 |
|  |  |  | Braunschweig | 25.04.2020 | 2 |
| North Rhine-Westphalia | 27.04.2020 | 27.04.2020 |  |  |  |
| Rheinland-Pfalz | 27.04.2020 | 27.04.2020 |  |  |  |
| Saarland | 27.04.2020 | 27.04.2020 |  |  |  |
| Saxony | 20.04.2020 | 20.04.2020 |  |  |  |
| Saxony-Anhalt | 22.04.2020 | 22.04.2020 |  |  |  |
| Schleswig-Holstein | 29.04.2020 | 29.04.2020 |  |  |  |
| Thuringia | 24.04.2020 | 24.04.2020 | Jena | 06.04.2020 | 18 |
|  |  |  | Nordhausen | 14.04.2020 | 10 |

*Notes:* A comprehensive overview of all public health measures introduced in German federal states and individual regions is given in Kleyer et al. (2020).

### B. Background and additional estimates for SCM application to Jena

This appendix presents supporting findings for the comparative case study of Jena.

#### B.1. Covid-19 cases and cumulative incidence rate in Jena and Germany on April 5

**Figure A1:**
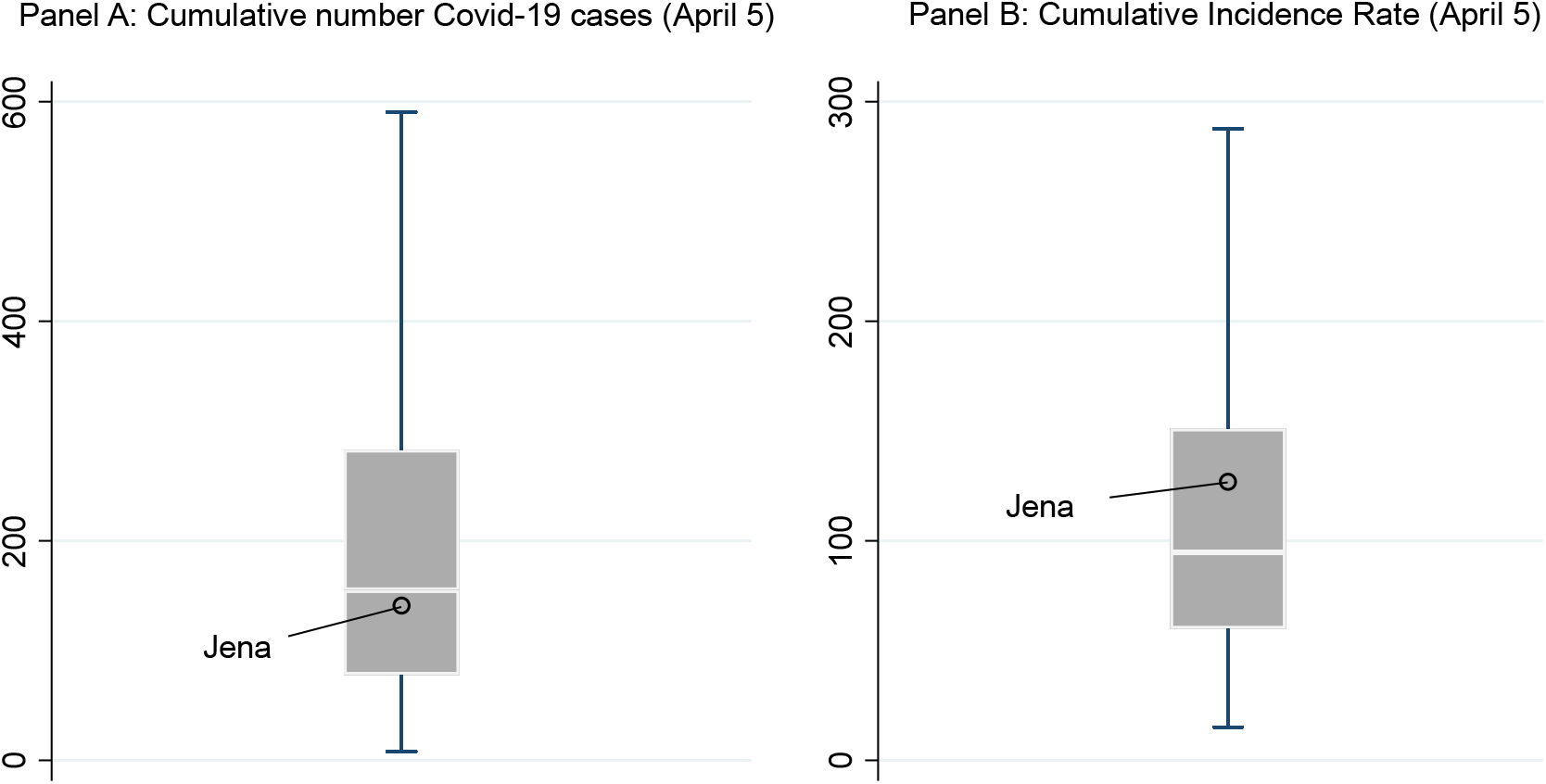
Box plots for distribution of Covid-19 cases across German NUTS3 regions (April 5)

#### B.2. Evaluation of pre-treatment predictor balance and prediction error (RMSPE)

This appendix shows the balancing properties of the SCM approach together with the root mean square percentage error (RMSPE) as a measure for the quality of the pre-treatment prediction.

**Table A2:**
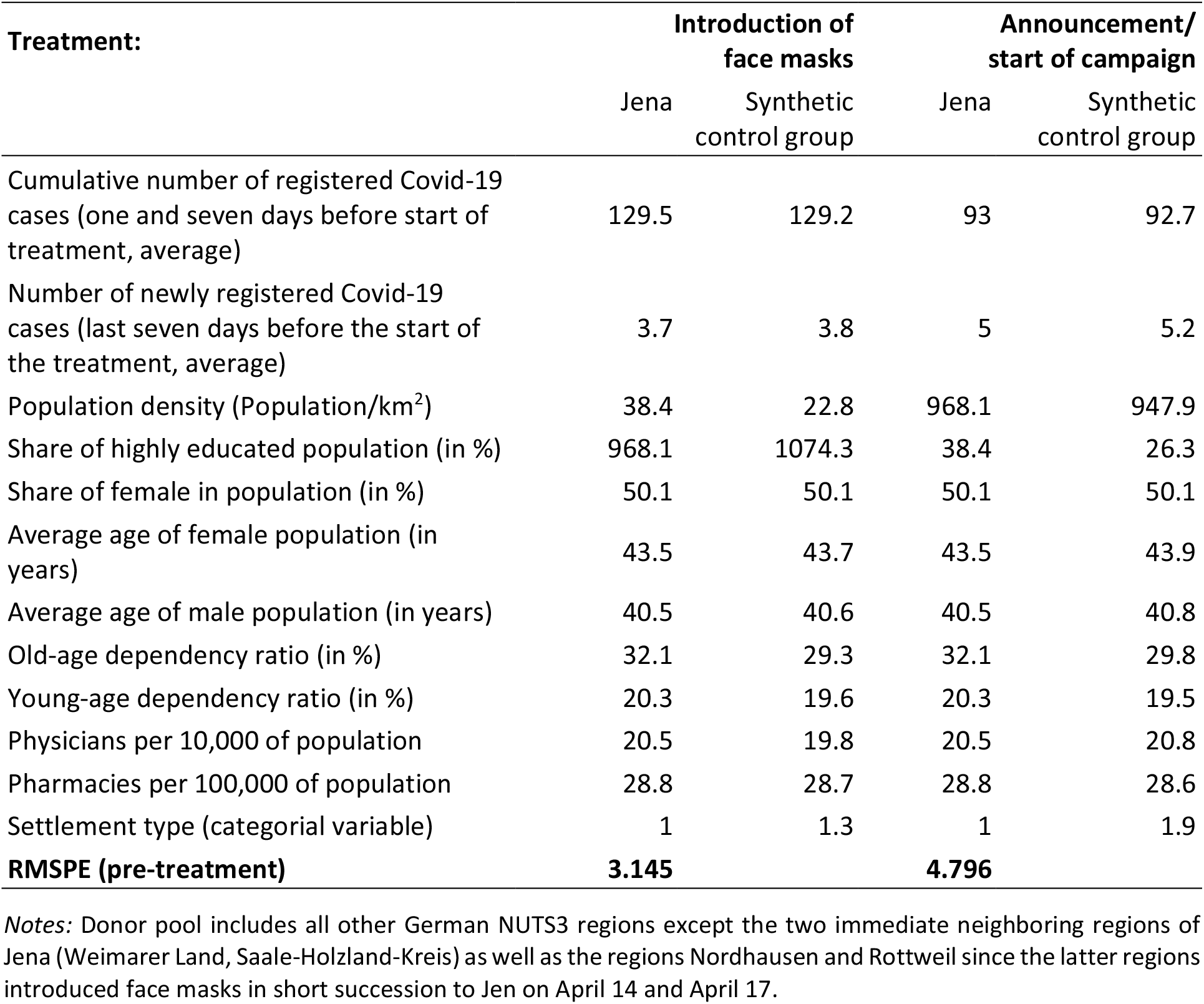
Pre-treatment predictor balance and RMSPE for SCM in Figure 2

#### B.3. Selected control regions and their associated sample weights

**Table A3:** Distribution of sample weights in donor pool for synthetic Jena

| Introduction of face masks (Panel A in Figure 2) |  |  |
| --- | --- | --- |
| ID | NUTS 3 region | Weight |
| 13003 | Rostock | 0.326 |
| 6411 | Darmstadt | 0.311 |
| 3453 | Cloppenburg | 0.118 |
| 7211 | Trier | 0.117 |
| 6611 | Kassel | 0.082 |
| 5370 | Heinsberg | 0.046 |

*Note:* Donor pools corresponds to SCM estimation in Panel A of Figure 2. Sample weights are chosen to minimize the RMSPE ten days prior to the start of the treatment.

#### B.4. Growth rates

Jena has 142 registered cases on April 6 compared to an estimated number of 143 cases in the synthetic control group. On April 26 Jena counts 158 cases and the synthetic control group reports 205 (again estimated) cases. The daily growth rate in Jena is denoted by x_Jena_ and can be computed from 142 [1+x_Jena_]^20^ = 158. The daily growth rate in the control group is denoted by x_control_ and can be computed from 143 [1+x_control_]^20^ = 205. Hence, the introduction of the face mask is associated with a decrease in the number of infections of x_control_ – x_Jena_ percentage points per day.

**Table A4:**
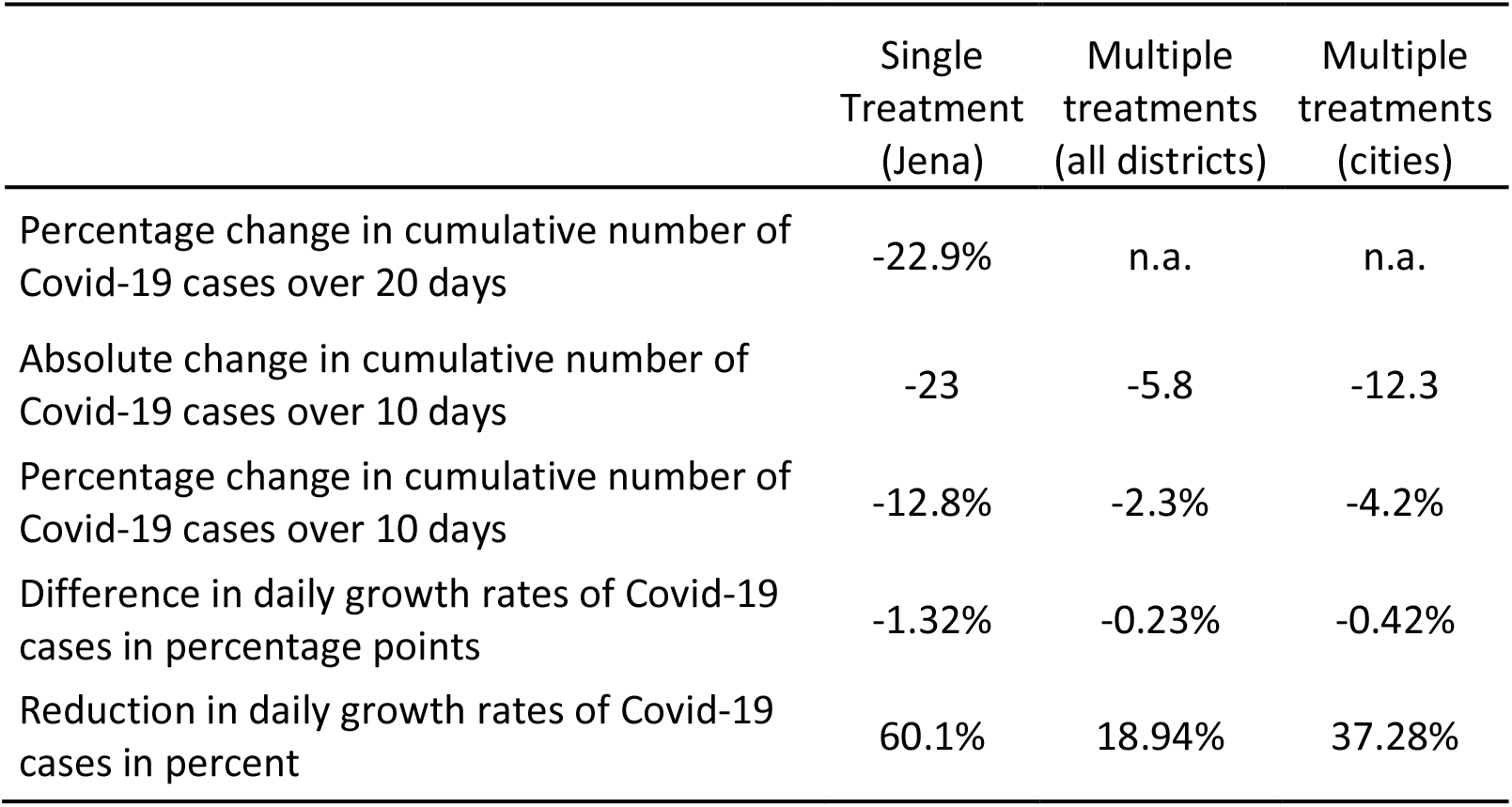
Summary of treatment effects of face mask introduction in Germany

These numbers are computed in an Excel-file available on the web pages of the authors.

#### B.5. SCM results by age groups

**Figure A2:**
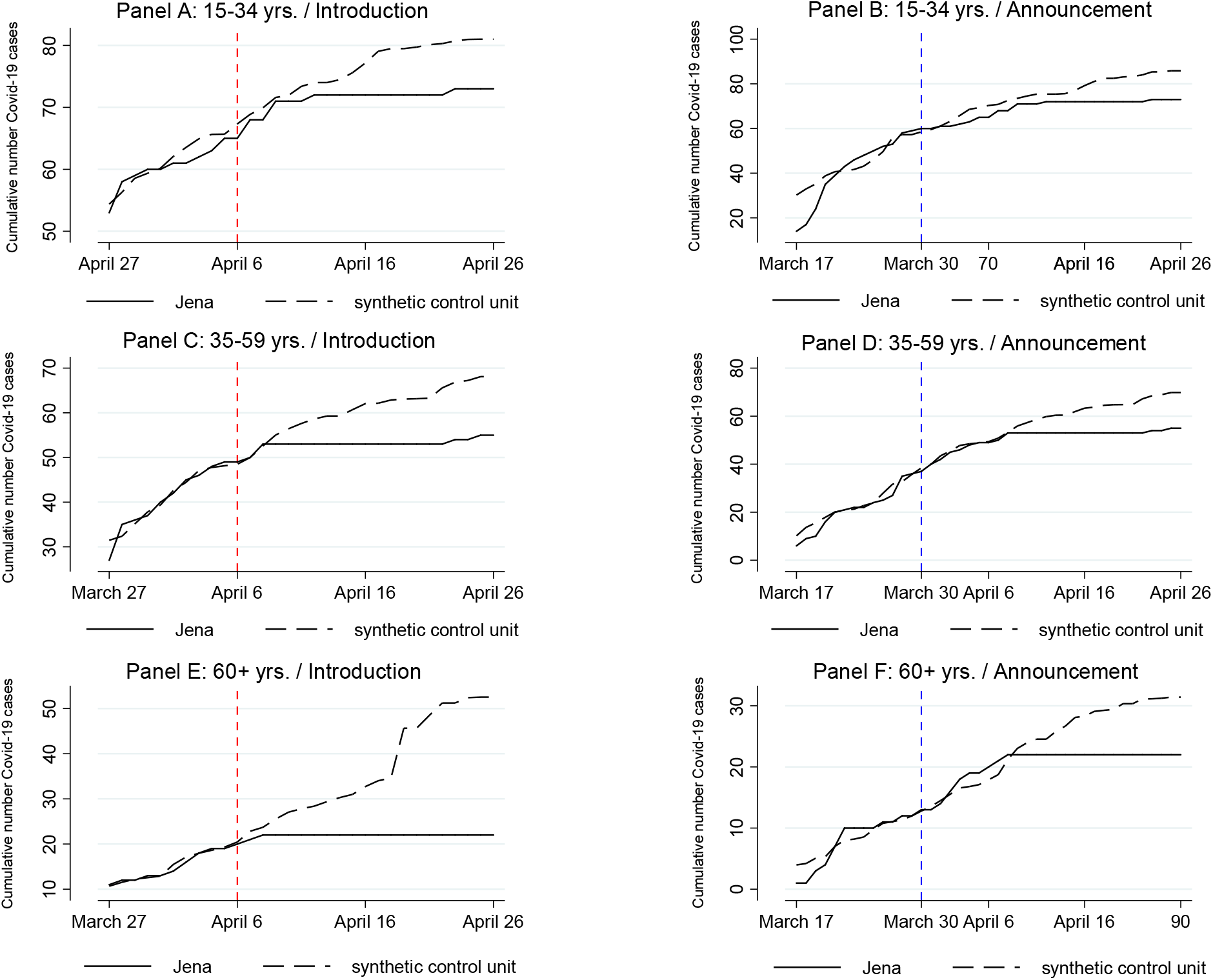
Treatment effects for introduction and announcement of face masks in Jena *Notes:* Predictor variables are chosen as for overall specification shown in Figure 2.

**Table A5:** Sample weights in donor pool for synthetic Jena (cumulative Covid-19 cases; by age groups)

| Age Group 15-34 years |  |  | Age Group 35-59 years |  |  | Age Group 60 years and above |  |  |
| --- | --- | --- | --- | --- | --- | --- | --- | --- |
| ID | NUTS 3 region | Weight | ID | NUTS 3 region | Weight | ID | NUTS 3 region | Weight |
| 1001 | Flensburg | 0.323 | 6411 | Darmstadt | 0.528 | 6411 | Darmstadt | 0.522 |
| 7211 | Trier | 0.207 | 16055 | Weimar | 0.16 | 16055 | Weimar | 0.244 |
| 13003 | Rostock | 0.184 | 14511 | Chemnitz | 0.15 | 7316 | Neustadt a.d. Weinstraße | 0.068 |
| 5370 | Heinsberg | 0.142 | 8221 | Baden-Baden | 0.07 | 9562 | Erlangen | 0.06 |
| 3453 | Cloppenburg | 0.107 | 6434 | Hochtaunus-kreis | 0.062 | 3356 | Osterholz | 0.056 |
| 6413 | Offenbach am Main | 0.038 | 8435 | Bodenseekreis | 0.029 | 5515 | Münster | 0.027 |
|  |  |  | 5370 | Heinsberg | 0.001 | 9188 | Starnberg | 0.022 |

Note: Donor pools corresponds to SCM estimations in Figure A2. Sample weights are chosen to minimize the RMSPE ten days prior to the start of the treatment.

#### B.6. Effects on cumulative number of infections per 100,000 inhabitants

**Figure A3:**
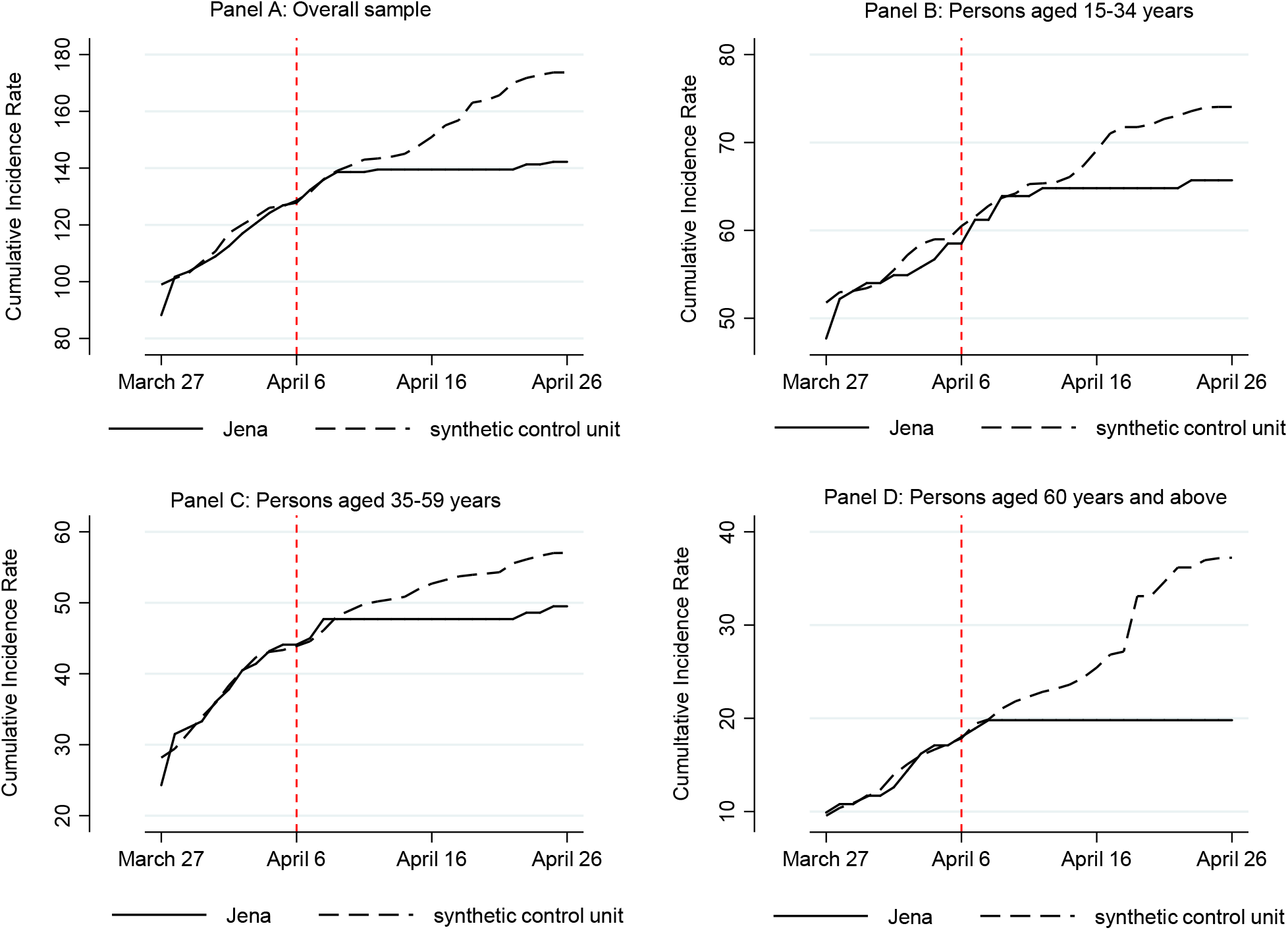
Treatment effects for introduction of face masks on cumulative incidence rate *Notes:* See Table 1 for a definition of the incidence rate. Predictor variables are chosen as for overall specification shown in Figure 2.

**Table A6:** Sample weights in donor pool for synthetic Jena (cumulative incidence rate)

| ID | NUTS 3 region | Weight |
| --- | --- | --- |
| 6411 | Darmstadt | 0.46 |
| 15003 | Magdeburg | 0.171 |
| 5370 | Heinsberg | 0.133 |
| 13003 | Rostock | 0.093 |
| 5515 | Münster | 0.066 |
| 11000 | Berlin | 0.035 |
| 12052 | Cottbus | 0.032 |
| 6611 | Kassel | 0.011 |

Note: Donor pools corresponds to SCM estimation in Figure A3. Sample weights are chosen to minimize the RMSPE ten days prior to the start of the treatment.

**Table A7:** Sample weights in donor pool for synthetic Jena (cumulative incidence rate; by age groups)

| Age Group 15-34 years |  |  | Age Group 35-59 years |  |  | Age Group 60 years and above |  |  |
| --- | --- | --- | --- | --- | --- | --- | --- | --- |
| ID | NUTS 3 region | Weight | ID | NUTS 3 region | Weight | ID | NUTS 3 region | Weight |
| 5370 | Heinsberg | 0.377 | 6411 | Darmstadt | 0.419 | 6411 | Darmstadt | 0.448 |
| 13003 | Rostock | 0.288 | 14511 | Chemnitz | 0.184 | 14612 | Dresden | 0.313 |
| 1001 | Flensburg | 0.14 | 14612 | Dresden | 0.154 | 9188 | Starnberg | 0.071 |
| 6611 | Kassel | 0.138 | 8221 | Heidelberg | 0.138 | 16054 | Suhl | 0.069 |
| 11000 | Berlin | 0.058 | 9188 | Starnberg | 0.088 | 5515 | Münster | 0.06 |
|  |  |  | 5370 | Heinsberg | 0.016 | 8221 | Heidelberg | 0.039 |

Note: Donor pools corresponds to SCM estimations in Figure A3. Sample weights are chosen to minimize the RMSPE ten days prior to the start of the treatment.

#### B.7. Google trends and announcement effects

**Figure A4:**
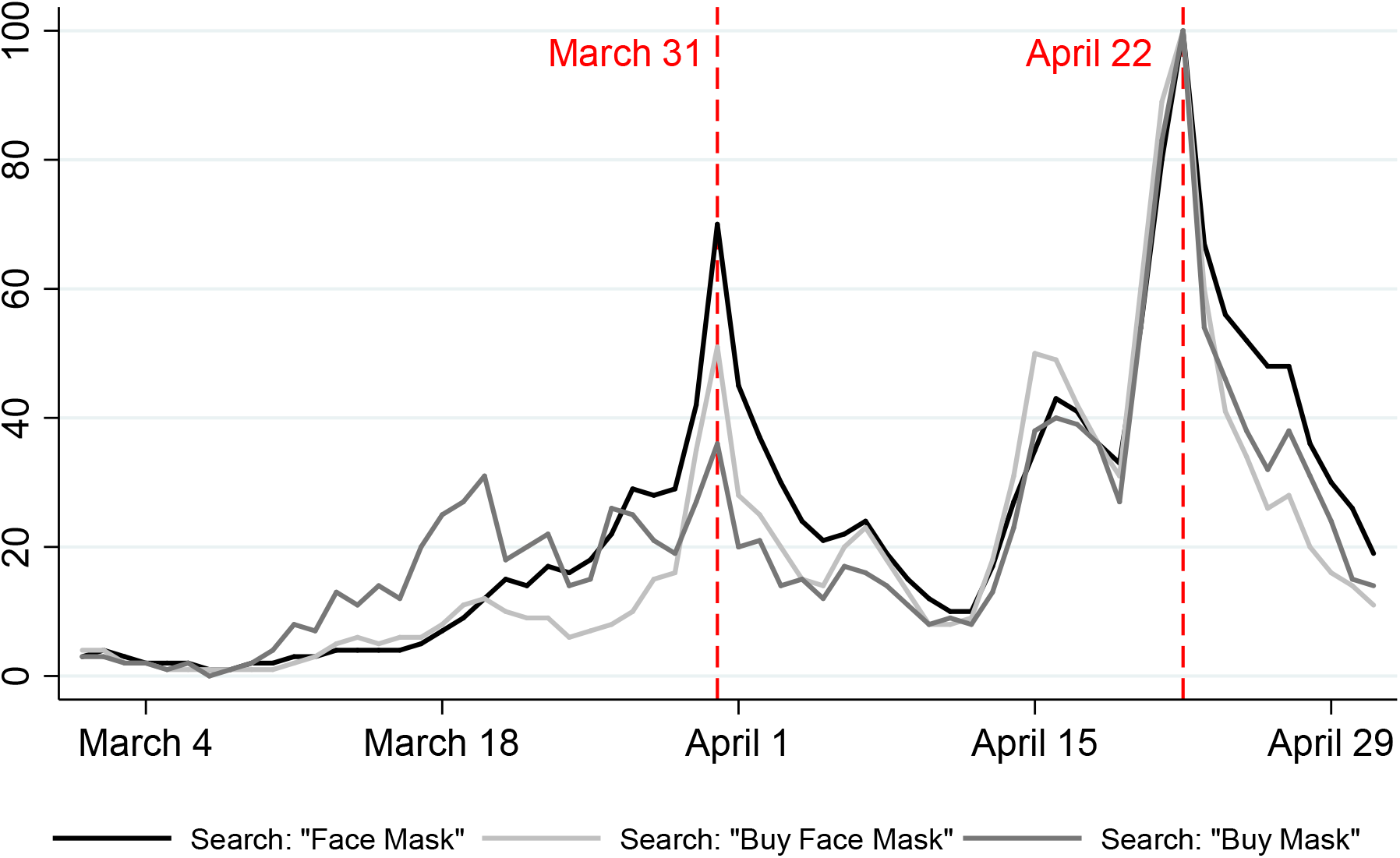
Online search for face masks and purchase options according to Google Trends *Note:* Online search for keywords (in German) as shown in the legend as Face Mask (“Mund.-Nasen-Schutz”), Buy Face Mask (“Mundschutz kaufen”) and Buy mask (“Maske kaufen”); alternative keywords show similar peaks but with a lower number of hits; based on data from Google Trends (2020).

#### B.8. Changes in donor pool for synthetic Jena

**Figure A5:**
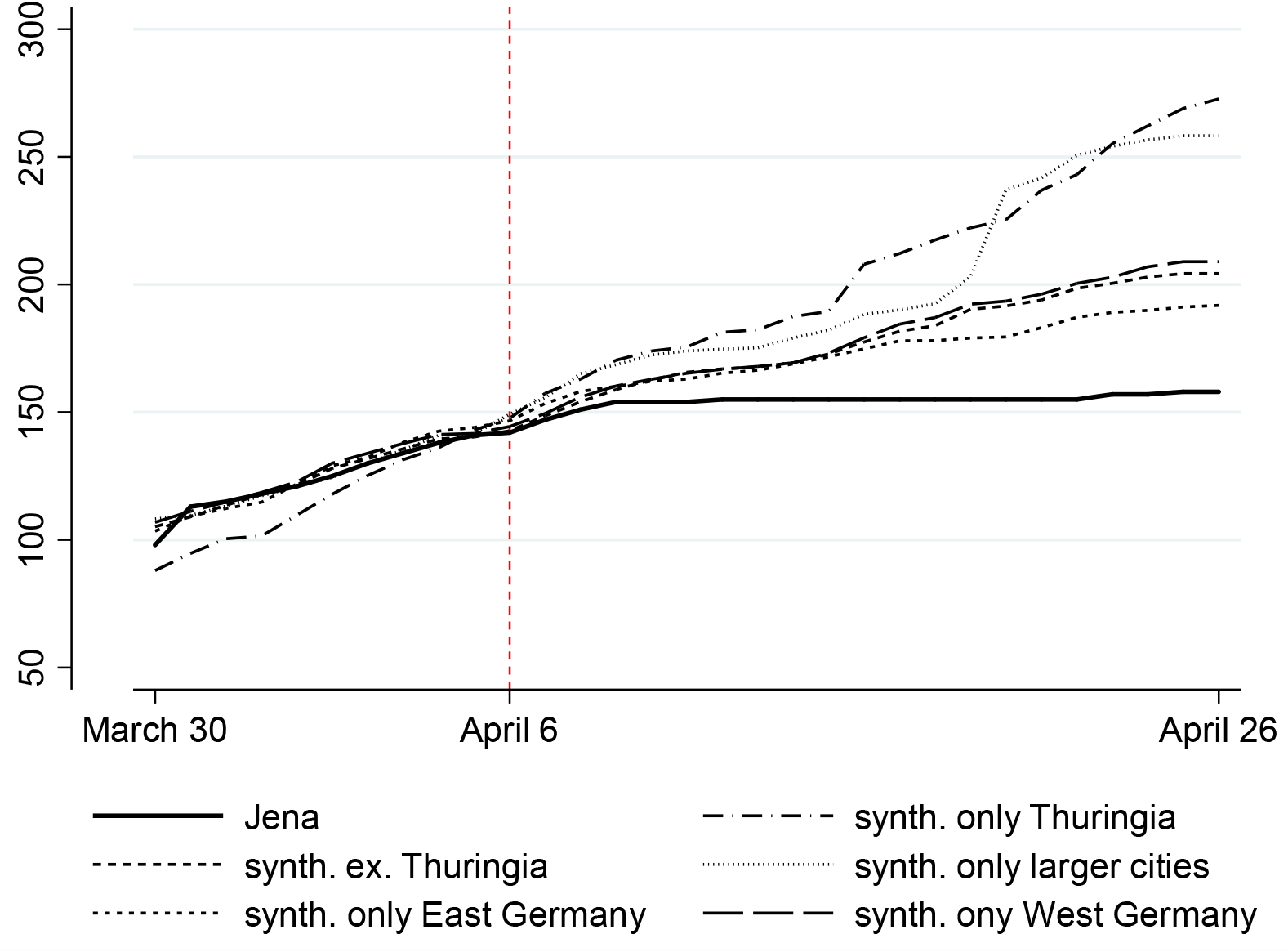
Treatment effects for changes in donor pool used to construct synthetic Jena *Notes:* See main text for a detailed definition of the respective donor pools. Predictor variables are chosen as for overall specification shown in Figure 2.

**Table A8:** Sample weights for alternative donor pools used to construct synthetic Jena

| Only Thuringia |  |  | Excluding Thuringia |  |  | Only larger cities |  |  |
| --- | --- | --- | --- | --- | --- | --- | --- | --- |
| ID | NUTS 3 region | Weight | ID | NUTS 3 region | Weight | ID | NUTS 3 region | Weight |
| 16076 | Greiz | 0.533 | 13003 | Rostock | 0.318 | 6411 | Darmstadt | 0.504 |
| 16051 | Erfurt | 0.467 | 6411 | Darmstadt | 0.302 | 13003 | Rostock | 0.304 |
|  |  |  | 7211 | Trier | 0.129 | 5113 | Essen | 0.192 |
|  |  |  | 3453 | Cloppenburg | 0.122 |  |  |  |
|  |  |  | 6611 | Kassel | 0.083 |  |  |  |
|  |  |  | 5370 | Heinsberg | 0.046 |  |  |  |
| Only East Germany |  |  | Only West Germany |  |  |  |  |  |
| ID | NUTS 3 region | Weight | ID | NUTS 3 region | Weight |  |  |  |
| 16051 | Erfurt | 0.865 | 6411 | Darmstadt | 0.242 |  |  |  |
| 14612 | Dresden | 0.124 | 3402 | Emden | 0.198 |  |  |  |
| 11000 | Berlin | 0.011 | 6611 | Kassel | 0.169 |  |  |  |
|  |  |  | 7211 | Trier | 0.168 |  |  |  |
|  |  |  | 4012 | Bremerhaven | 0.167 |  |  |  |
|  |  |  | 5370 | Heinsberg | 0.057 |  |  |  |

Note: Donor pools corresponds to SCM estimations in Figure A5. Sample weights are chosen to minimize the RMSPE ten days prior to the start of the treatment.

#### B.9. Place-in-space tests for other major cities in Thuringia

**Figure A6:**
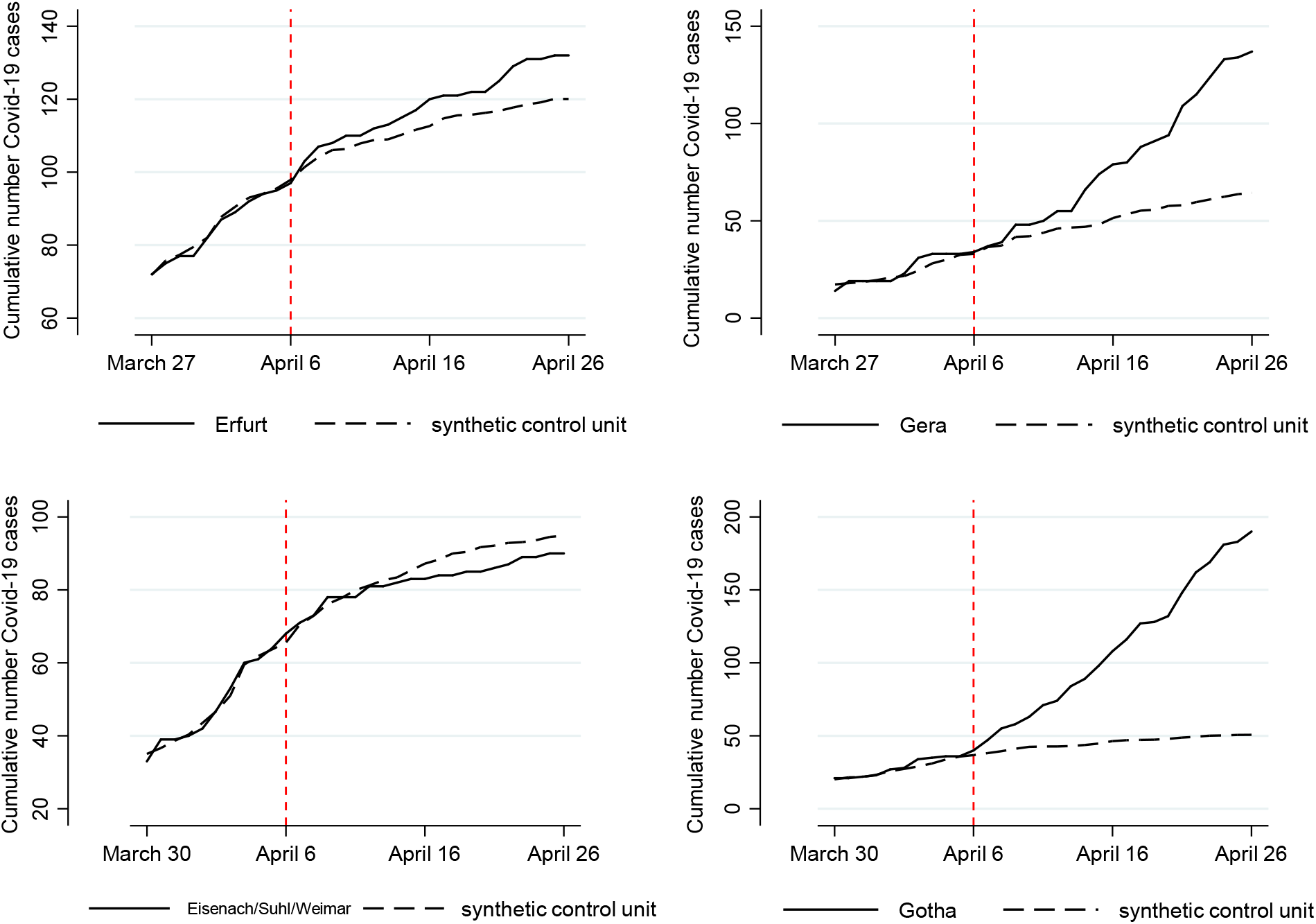
Placebo tests for the effect of face masks in other cities in Thuringia on April 6. *Notes:* For the placebo tests in the other cities in Thuringia the same set of predictors as for Jena (Figure 2) has been applied. The reported regions cover all *kreisfreie Städte* plus Gotha (*Landkreis*). The cities Weimar, Suhl and Eisenach have been aggregated since the number of reported Covid-19 is low in these cities.

**Table A9:**
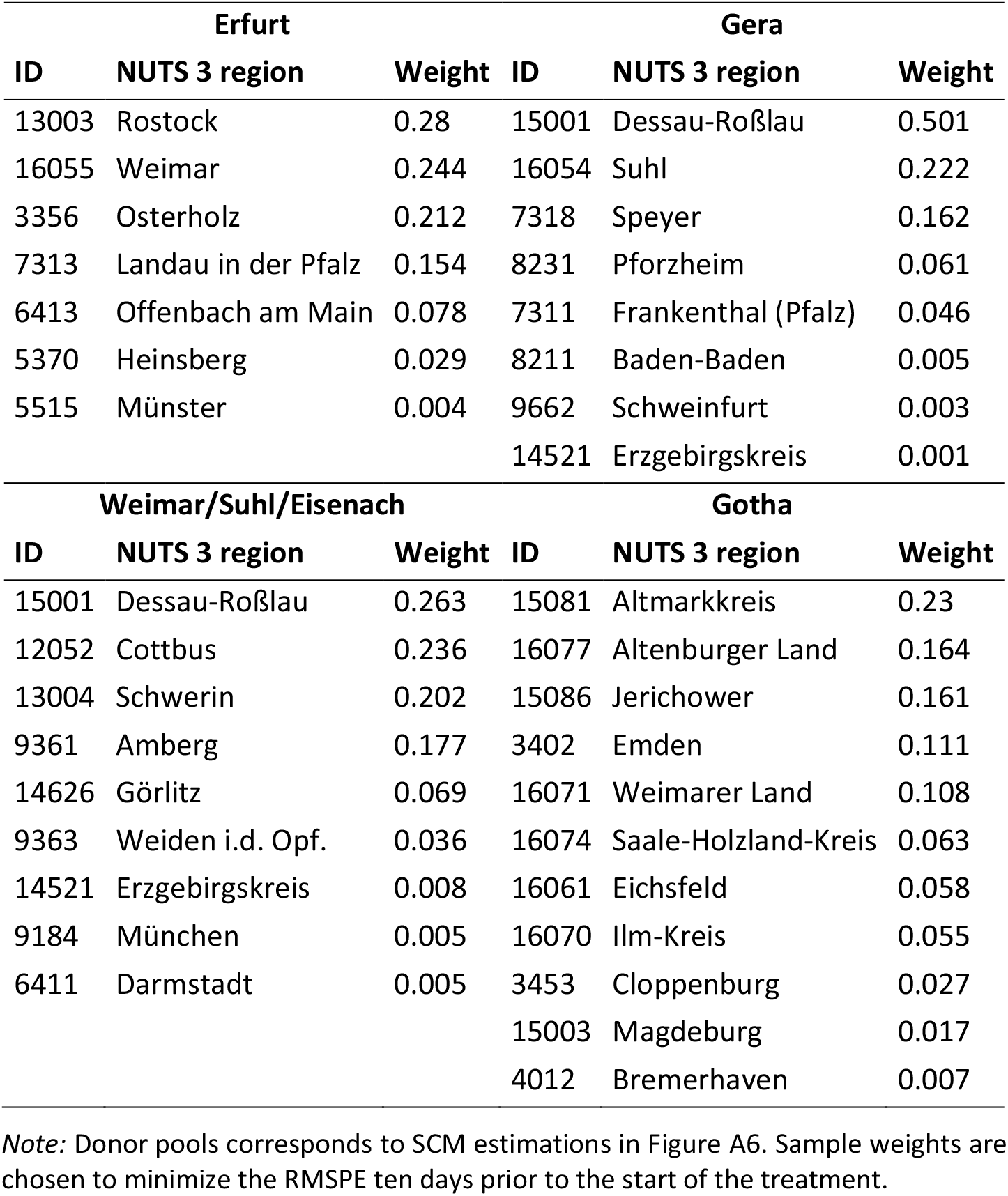
Sample weights in donor pool for synthetic control groups (other cities in Thuringia)

| Erfurt |  |  | Gera |  |  |
| --- | --- | --- | --- | --- | --- |
| ID | NUTS 3 region | Weight | ID | NUTS 3 region | Weight |
| 13003 | Rostock | 0.28 | 15001 | Dessau-Roßlau | 0.501 |
| 16055 | Weimar | 0.244 | 16054 | Suhl | 0.222 |
| 3356 | Osterholz | 0.212 | 7318 | Speyer | 0.162 |
| 7313 | Landau in der Pfalz | 0.154 | 8231 | Pforzheim | 0.061 |
| 6413 | Offenbach am Main | 0.078 | 7311 | Frankenthal (Pfalz) | 0.046 |
| 5370 | Heinsberg | 0.029 | 8211 | Baden-Baden | 0.005 |
| 5515 | Münster | 0.004 | 9662 | Schweinfurt | 0.003 |
|  |  |  | 14521 | Erzgebirgskreis | 0.001 |
| Weimar/Suhl/Eisenach |  |  | Gotha |  |  |
| ID | NUTS 3 region | Weight | ID | NUTS 3 region | Weight |
| 15001 | Dessau-Roßlau | 0.263 | 15081 | Altmarkkreis | 0.23 |
| 12052 | Cottbus | 0.236 | 16077 | Altenburger Land | 0.164 |
| 13004 | Schwerin | 0.202 | 15086 | Jerichower | 0.161 |
| 9361 | Amberg | 0.177 | 3402 | Emden | 0.111 |
| 14626 | Görlitz | 0.069 | 16071 | Weimarer Land | 0.108 |
| 9363 | Weiden i.d. Opf. | 0.036 | 16074 | Saale-Holzland-Kreis | 0.063 |
| 14521 | Erzgebirgskreis | 0.008 | 16061 | Eichsfeld | 0.058 |
| 9184 | München | 0.005 | 16070 | Ilm-Kreis | 0.055 |
| 6411 | Darmstadt | 0.005 | 3453 | Cloppenburg | 0.027 |
|  |  |  | 15003 | Magdeburg | 0.017 |
|  |  |  | 4012 | Bremerhaven | 0.007 |

*Note:* Donor pools corresponds to SCM estimations in Figure A6. Sample weights are chosen to minimize the RMSPE ten days prior to the start of the treatment.

### C. The effect in other German cities and regions (single treatment analyses)

In addition to Jena, we test for treatment effects in Nordhausen, Rottweil, Main-Kinzig-Kreis, and Wolfsburg (compare Figure 1). We ignore Braunschweig here as the introduction became effective only two days in advance of its federal state.

**Figure A7:**
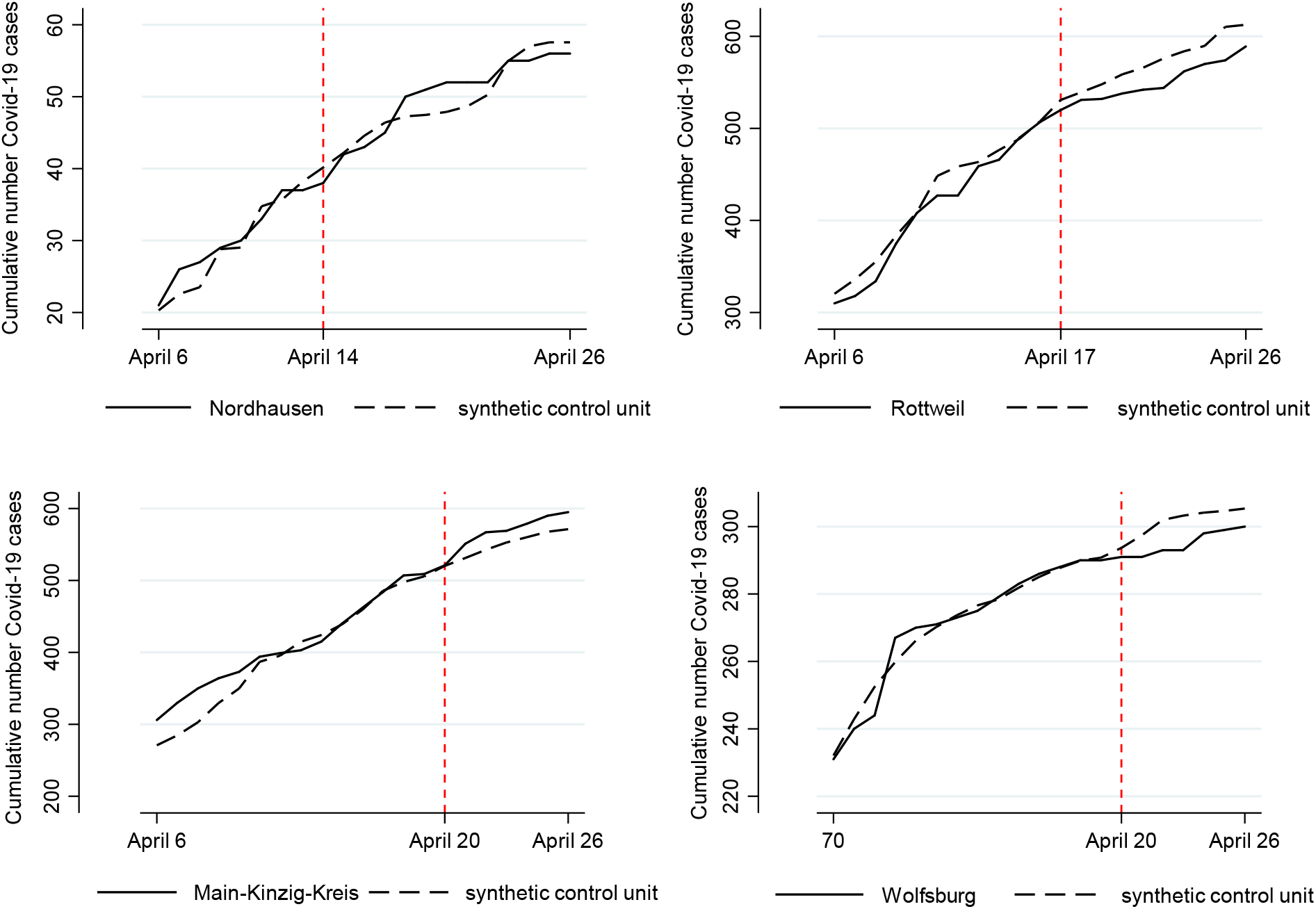
Treatment effects for introduction of face masks in other cities *Notes:* Nordhausen (Thuringia, April 14, top left), Rottweil (Baden Württemberg, April 17, top right), Wolfsburg (Lower Saxony, April 20, middle left), Main-Kinzig-Kreis (Hessia, April 20, middle right). Predictor variables are chosen as for overall specification shown in Figure 2.

As the figure shows, the result is 2:1:1. Rottweil and Wolfsburg display a positive effect of mandatory mask wearing, just as Jena. The results in Nordhausen are very small or unclear. In the region of Main-Kinzig, it even seems to be the case that masks increased the number of cases relative to the synthetic control group. As all of these regions introduced masks after Jena, the time period available to identify effects is smaller than for Jena. The effects of mandatory face masks could also be underestimated as announcement effects and learning from Jena might have induced individuals to wear masks already before they became mandatory. Finally, the average pre-treatment RMSPE for these four regions (7.150) is larger than for the case of Jena (3.145). For instance, in the case of the region of Main-Kinzig it is more than three times as high (9.719), which indicates a lower pre-treatment fit. The obtained treatment effects should then be interpreted with some care as the pre-sample error could also translate into the treatment period. In order to minimize the influence of a poor pre-treatment fit for some individual regions, the main text therefore compares the results in Jena mainly with a multiple unit treatment approach.

**Table A10:**
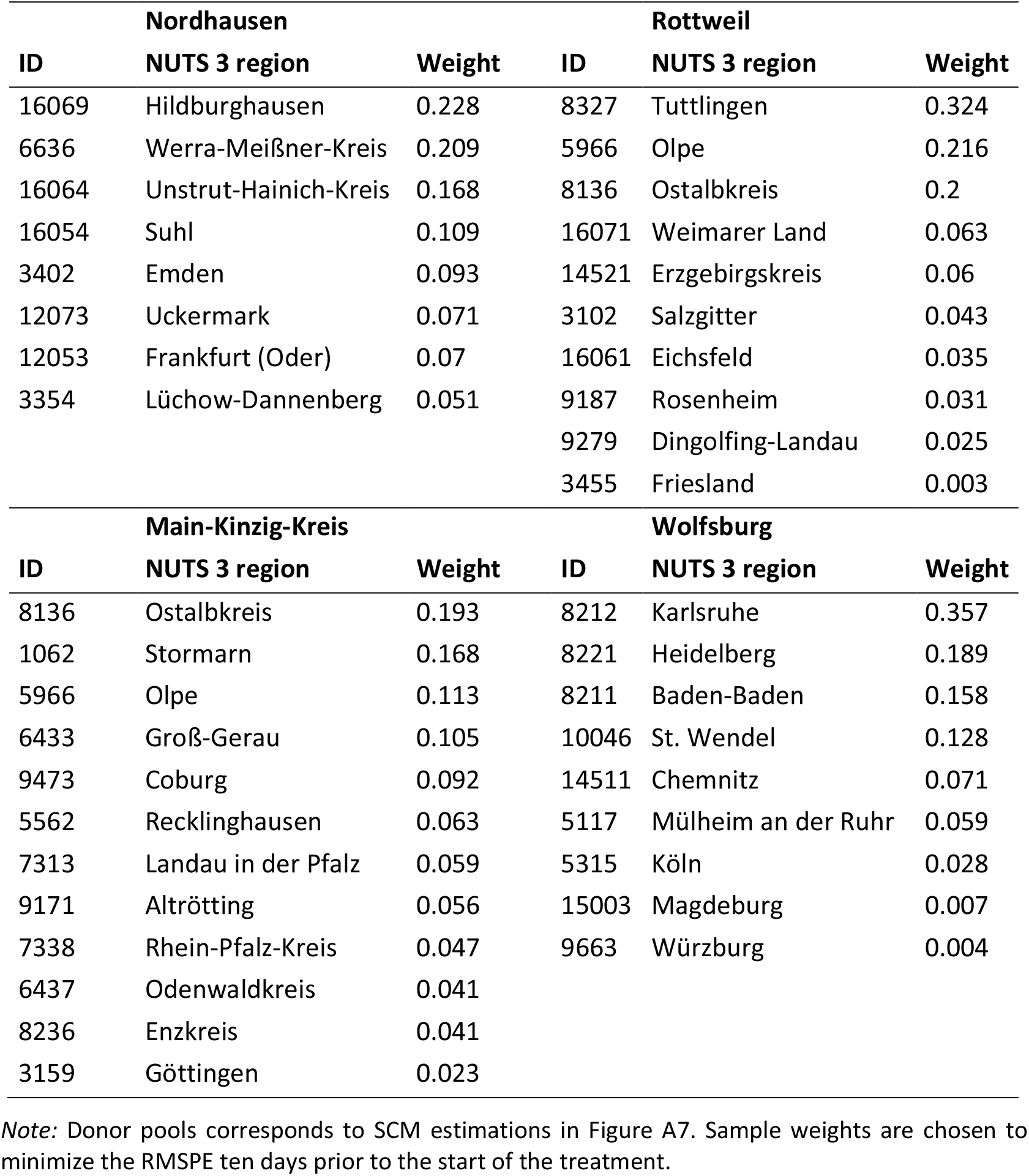
Sample weights in donor pool for synthetic controls (other treated NUTS3 regions)

| **Nordhausen** |  |  | **Rottweil** |  |  |
| --- | --- | --- | --- | --- | --- |
| **ID** | **NUTS 3 region** | **Weight** | **ID** | **NUTS 3 region** | **Weight** |
| 16069 | Hildburghausen | 0.228 | 8327 | Tuttlingen | 0.324 |
| 6636 | Werra-Meißner-Kreis | 0.209 | 5966 | Olpe | 0.216 |
| 16064 | Unstrut-Hainich-Kreis | 0.168 | 8136 | Ostalbkreis | 0.2 |
| 16054 | Suhl | 0.109 | 16071 | Weimarer Land | 0.063 |
| 3402 | Emden | 0.093 | 14521 | Erzgebirgskreis | 0.06 |
| 12073 | Uckermark | 0.071 | 3102 | Salzgitter | 0.043 |
| 12053 | Frankfurt (Oder) | 0.07 | 16061 | Eichsfeld | 0.035 |
| 3354 | Lüchow-Dannenberg | 0.051 | 9187 | Rosenheim | 0.031 |
|  |  |  | 9279 | Dingolfing-Landau | 0.025 |
|  |  |  | 3455 | Friesland | 0.003 |
| **Main-Kinzig-Kreis** |  |  | **Wolfsburg** |  |  |
| **ID** | **NUTS 3 region** | **Weight** | **ID** | **NUTS 3 region** | **Weight** |
| 8136 | Ostalbkreis | 0.193 | 8212 | Karlsruhe | 0.357 |
| 1062 | Stormarn | 0.168 | 8221 | Heidelberg | 0.189 |
| 5966 | Olpe | 0.113 | 8211 | Baden-Baden | 0.158 |
| 6433 | Groß-Gerau | 0.105 | 10046 | St. Wendel | 0.128 |
| 9473 | Coburg | 0.092 | 14511 | Chemnitz | 0.071 |
| 5562 | Recklinghausen | 0.063 | 5117 | Mülheim an der Ruhr | 0.059 |
| 7313 | Landau in der Pfalz | 0.059 | 5315 | Köln | 0.028 |
| 9171 | Altrötting | 0.056 | 15003 | Magdeburg | 0.007 |
| 7338 | Rhein-Pfalz-Kreis | 0.047 | 9663 | Würzburg | 0.004 |
| 6437 | Odenwaldkreis | 0.041 |  |  |  |
| 8236 | Enzkreis | 0.041 |  |  |  |
| 3159 | Göttingen | 0.023 |  |  |  |

*Note:* Donor pools corresponds to SCM estimations in Figure A7. Sample weights are chosen to minimize the RMSPE ten days prior to the start of the treatment.

### D. A brief survey of public health measures against Covid-19

Our approach goes in line with various studies that have already tried to better understand the effect of public health measures on the spread of Covid-19 (Barbarossa et al., 2020, Hartl et al., 2020, Donsimoni et al., 2020, Dehning et al., 2020, Gros et al., 2020, Adamik et al, 2020). However, these earlier studies all take an aggregate approach in the sense that they look at implementation dates for a certain measure and search for subsequent changes in the national incidence. There are some prior analyses that take a regional focus (Khailaie et al. 2020) but no attention is paid to the effect of policy measures.^11^

There are also many cross-country analyses, both in a structural SIR (susceptible, infectious and removed) sense (Chen and Qiu, 2020) and with an econometric focus on forecasting the end of the pandemic (Ritschl, 2020). Others draw parallels between earlier pandemics and Covid-19 (Barro et al., 2020). These studies do not explicitly take public health measures into account. Some studies discuss potential effects of public health measures and survey general findings (Wilder-Smith et al. 2020, Anderson et al., 2020, Ferguson et al, 2020) but do not provide direct statistical evidence on specific measures.

The synthetic control method (SCM) has been applied by Friedson et al. (2020) to estimate the effect of the shelter-in-place order for California, USA, in the development of Covid-19. The authors find *inter alia* that around 1600 deaths from Covid-19 have been avoided by this measure during the first four weeks. The effects of face masks have been surveyed by Howard et al. (2020) and Greenhalgh et al. (2020). Greenhalgh et al. (2020) mainly presents evidence on the effect of face masks during non-Covid epidemics (influenza and SARS). Marasinghe (2020) reports that they “*did not find any studies that investigated the effectiveness of face mask use in limiting the spread of COVID-19 among those who are not medically diagnosed with COVID-19 to support current public health recommendations*”.

In addition to medical aspects (like transmission characteristics of Covid-19 and filtering capabilities of masks), Howard et al. (2020) survey evidence on mask efficiency and on the effect of a population. They first stress that “*no randomized control trials on the use of masks <…> has been published*”. The study which is “*the most relevant paper*” for Howard et al. (2020) is one that analyzed *“exhaled breath and coughs of children and adults with acute respiratory illness*” (Leung et al., 2020, p. 676), i.e. used a clinical setting. Concerning the effect of masks on community transmissions, the survey needs to rely on pre-Covid-19 studies.

We conclude from this literature review that our paper is the first analysis that provides field evidence on the effect of masks on mitigating the spread of Covid-19.

## Footnotes

2 This is similar to the setup in Abadie et al. (2010), who study the effect of an increase in the tobacco tax in California. The tobacco tax was decided upon by the state government.

3 The main channel through which masks reduce transmission of SARS-CoV-2 is the reduction in aerosols and droplets, as argued by Prather et al. (2020).

4 Friedson et al. (2020) employ the SCM to estimate the effect of the shelter-in-place order for California in the development of Covid-19. The authors find *inter alia* that around 1600 deaths from Covid-19 were avoided by this measure during the first four weeks.

5 We are aware of the existence of hidden infections. As it appears plausible to assume that they are proportional to observed infections across regions, we do not believe that they affect our results. We chose the date of reporting (as opposed to date of infections) because not all reported infections include information about the date of infection.

6 We conduct all estimations in STATA using “Synth” and “Synth Runner” packages (Abadie at al., 2020, Galiani and Quistorff, 2017). Data and estimation files can be obtained from the authors upon request.

7 The pre-treatment root mean square prediction error (RMSPE) of 3.145 is significantly below a benchmark RMSPE of 6.669, which has been calculated as the average RMSPE for all 401 regions in the pre-treatment period until April 6. This points to the relatively good fit of the synthetic control group for Jena in this period.

8 See https://www.tagesschau.de/inland/corona-maskenpflicht-103.html. Last accessed May 05, 2020.

9 See https://www.jenaer-nachrichten.de/stadtleben/13069-jena-zeigt-maske-kampagne-f%C3%BCr-mund-schutz-startet. Last accessed May 05, 2020.

10 This is perfectly in line with Prather et al. (2020) given the reduction in aerosols and droplets via using masks.

11 In a short note, Hartl and Weber (2020) apply panel methods based on time dummies to understand the relative importance of various public health measures. They employ data at the federal state level and not at the regional level. As a detailed model description is not available, an appreciation of results is difficult at this point.

